# The Evaluation of Tenecteplase for the Treatment of an Ischemic Stroke

**DOI:** 10.1101/2025.11.05.25339636

**Authors:** William Braun, Allan Weiss, Harold Colbassani, Ajay Arora, Paul Lewis, Eric Lopez del Valle, Marissa Lepain, Christi Newman, Keith Chastain

**Author notes:** The authors declare no relevant or material financial interests that relate to the research described in this paper. All authors have read and approved the submitted manuscript, the manuscript has not been submitted elsewhere nor published elsewhere in whole or in part, except as an abstract (if relevant).

## Abstract

**Introduction:** For more than two decades, the standard for thrombolysis for Acute Ischemic Stroke (AIS) has been alteplase 0.9 mg/kg (maximum 90 mg) with 10 % given as a bolus. More recent studies such as the EXTEND-IA, and AcT study have shown tenecteplase (TNK) may have potential advantages over alteplase for AIS. In November 2024 the ATTEST-2 study was published with the largest comparative prospective study to date. This study included 1858 patients from 39 stroke centers in the UK which were randomized to receive TNK 0.25 mg/kg IV or alteplase standard of care dose. The study showed TNK was non-inferior to alteplase for 90-day mRS (p<0.001). On March 3^rd^, 2025, the FDA approved TNK for AIS based off the AcT trial. This study further evaluates the effectiveness and safety of TNK at 0.25 mg/kg IV for AIS and represents real world community hospital utilization covering a diverse population reinforcing the safety and efficacy of TNK compared with alteplase.

**Methods:** The analysis used a multicenter, retrospective cohort study across 11 primary and comprehensive hospitals within the BayCare Health System in West Central Florida (December 2019–April 2024), comparing TNK 0.25 mg/kg (max 25 mg) with standard-dose alteplase among adults with AIS presenting within 4.5 hours of last-known-well. The primary outcome (LVO subgroup) was substantial early reperfusion prior to thrombectomy (mTICI 2b/2c/3 or absence of retrievable thrombus). Key secondary outcomes included door-to-needle time, admission, and discharge NIHSS and 90-day mRS, and safety (ICH, angioedema). Adjusted analyses used logistic and ordinal logistic regression.

**Results:** Among 476 AIS patients (TNK n=270; alteplase n=206), 226 had LVO (TNK n=115; alteplase n=111). Early reperfusion occurred in 14.8% with TNK vs 4.5% with alteplase (risk difference 10.3% [95% CI, 2.7–17.8], p=0.008). Adjusted odds of early reperfusion were higher with TNK (OR 3.53 [95% CI, 1.20–10.40], p=0.022). In all AIS patients, median door-to-needle time was shorter with TNK (34 vs 45 minutes; difference −11 [95% CI, −14.4 to −7.6], p<0.001). Good 90-day functional outcome (mRS 0–2) was more common with TNK among LVO patients (47.3% vs 29.3%, p=0.031) and among all AIS patients (61.8% vs 50.0%, difference 11.8% [95% CI, −0.3 to 23.4], p=0.044). Symptomatic ICH and angioedema were similar; across all patients combined, ICH (symptomatic and asymptomatic) was less frequent with TNK (8.5% vs 15.0%, p=0.030).

**Conclusion:** In routine practice, TNK 0.25 mg/kg was associated with higher pre-thrombectomy reperfusion in LVO, shorter door-to-needle times, comparable safety, and improved functional outcomes versus alteplase in key analyses. These findings align with recent randomized evidence and support TNK as an effective alternative to alteplase for AIS.

## Introduction

For more than twenty years since Food and Drug Administration (FDA) approval in 1996, alteplase 0.9 mg/kg (maximum dose 90 mg with 10 % given as a bolus) has been the standard of care for thrombolysis in the setting of Acute Ischemic Stroke (AIS). TNK was engineered by modifying the Alteplase structure using point mutation of the Alteplase 527 amino acid glycoprotein structure at three sites, threonine 103, asparagine 117 and kringle 1 domain. These modifications resulted in improved specificity to fibrin and being more resistant to degradation by endogenous enzymes plasminogen activator inhibitor-1(PAI-1) resulting in longer half-life.^17^

With these modifications, TNK has faster rate of administration, greater efficacy, lower bleed risk, and faster door-to-needle time (DTN). In June 2000 TNK was FDA approved for ST-segment elevated myocardial infarction (STEMI) as a 5 to 10-minute intravenous push dose compared to Alteplase 60-minute infusion. This was based on the results from the ASSENT-2 trial which showed TNK and Alteplase when used for STEMI had similar reductions in 30-day mortality and rates of intracranial hemorrhages.^18^ More recent studies have shown that TNK may have potential advantages over Alteplase for AIS. There have been several large, randomized trials evaluating TNK for the treatment of AIS. The NOR-TEST study was one the first large prospective studies with 1100 patients in combined study arms.^5^ The trial demonstrated TNK had favorable 90 day functional outcomes compared to Alteplase and showed similar safety and efficacy.^5^ Burgos et al. conducted a meta-analysis of five randomized trials comparing Alteplase to TNK for AIS which showed TNK to be non-inferior to alteplase for the treatment of AIS with similar Intracerebral hemorrhage (ICH) rates with 3% in both groups.^11^ The EXTEND-IA TNK study showed that TNK 0.25 mg/kg IV given before thrombectomy was associated with higher rate of reperfusion and better functional outcomes than the standard dose of Alteplase in AIS patients with a large vessel occlusion (LVO), if treated within 4.5 hrs. of stroke onset.^3^ The EXTEND-IA TNK part 2 showed that higher TNK doses (0.40 mg/kg) compared with 0.25 mg/kg had no difference in improving cerebral reperfusion prior to endovascular thrombectomy. However, symptomatic ICH occurred in seven patients (4.7%) in the 0.40 mg/kg group compared to two (1.3%) in the 0.25 mg/kg group.^12^ Currently the 2019 American Heart Association/ American Stroke Association (AHA/ASA) guidelines give a IIb recommendation for choosing TNK over alteplase in patients without contraindications for IV fibrinolysis who are eligible to undergo mechanical thrombectomy.^25^ In 2022, the AcT trial studied 1577 patients at 22 centers across Canada. It compared TNK 0.25 mg/kg (max 25 mg) to standard alteplase dosing for AIS within 4.5hrs.^20^ The Act trial showed TNK to be non-inferior to alteplase for the treatment of AIS with primary outcome Modified Rankin Score (mRS) 0-1 was 36.9% in TNK group and 34.8% in the alteplase group with similar safety outcomes.^20^ In November 2024 the ATTEST-2 study was published with the largest comparative prospective study to date. This study included 1858 patients from 39 stroke centers in the UK which were randomized to receive TNK 0.25 mg/kg or alteplase standard of care dose. The study showed TNK was non-inferior to alteplase for 90-day mRS. The study did not demonstrate that TNK was superior for 90-day mRS distribution compared to alteplase. Symptomatic ICH and angioedema were similar between the groups.^19^ On March 3^rd^, 2025, the Food and Drug Administration (FDA) approved TNK for AIS based off the AcT trial.^27^ The aim of this study was to retrospectively compare alteplase standard dosing to TNK 0.25mg/kg Max 25 mg for AIS in clinical practice for patients with LVO and all patients presenting with AIS.

## Methods

### Study Design and Setting

This multicenter, retrospective cohort study examined a multicenter, observational, retrospective cohort study across 11 primary and comprehensive stroke centers in a single health system in West-Central Florida. Consecutive adults (≥18 years) with AIS treated with intravenous TNK 0.25 mg/kg (max 25 mg) or standard-dose alteplase (0.9 mg/kg, max 90 mg) between December 2019 and April 2024 were identified from the electronic health record (Cerner PowerChart). Patients presented within 4.5 hours of last-known-well and met system criteria for thrombolysis.

### Outcomes

The primary outcome (LVO subgroup) was substantial early reperfusion prior to thrombectomy, defined as restoration of blood flow to >50% of the ischemic territory (mTICI 2b/2c/3) by CTA/MRA or absence of a retrievable thrombus at the initial angiographic assessment. A large vessel occlusion (LVO) was determined at the initial CT/CTA prior to the thrombectomy. Secondary outcomes included door-to-needle time (DTN), NIHSS at 24 hours, discharge NIHSS, discharge disposition, 90-day mRS, and safety (symptomatic/asymptomatic ICH by system definitions and angioedema). Safety outcomes for ICH were assessed using the SITS-MOST criteria for symptomatic ICH including subarachnoid hemorrhage within 36 hours of treatment combined with an increase from baseline in NIHSS score of at least 4 points.^28^

### Predictive Margins

To aid interpretability of adjusted models, estimates were obtained using predictive margins in Stata 15. For each outcome, predicted probabilities were computed adjusted, model-based probabilities by treatment group across clinically relevant ranges of age (e.g., deciles spanning middle to advanced age) and initial NIHSS (across the observed distribution). All other covariates were held at their observed values, so marginal predictions reflect the sample’s covariate distribution. Point estimates are shown with 95% CIs. These margins complement odds ratios by translating covariate effects into predicted probabilities.

### Statistical Analysis

Continuous variables were summarized as means with standard deviations or medians with interquartile ranges, as appropriate; categorical variables as counts and percentages. Two-sample t-tests compared means; quantile (median) regression compared non-normal continuous and ordinal variables; and two-proportion z-tests (or Fisher’s exact tests when expected counts <5) compared proportions. Multivariable logistic regression estimated adjusted odds ratios (ORs) and 95% confidence intervals (CIs) for binary outcomes (e.g., early reperfusion, 90-day mRS 0–1, 0–2, mortality), and proportional-odds ordinal logistic regression modeled mRS distributions at discharge and 90 days. Models adjusted for treatment group and prespecified covariates (age per 10 years, initial NIHSS, history of cerebrovascular disease, hypertension, atrial fibrillation, diabetes, cardiovascular disease, hyperlipidemia, smoking). A two-sided α of 0.05 defined statistical significance.

The analysis used Minitab 22 for descriptives and two-proportion tests, and Stata 15 for binary logistic, ordinal logistic and quantile regressions).

Sample size calculation was based on the EXTEND-IA TNK trial. Reperfusion occurred in 22% of the TNK group compared to 10% in the alteplase group with an incidence difference of 12%. Based on this study it was estimated that sample size of 200 would achieve 90% power for detecting a 10% difference. A 10% overestimation was added to the 200-sample size estimation to ensure 80% power was obtained. The primary analysis was based on the intention to treat population, and any missing data remained missing. All hypothesis tests used a 2-sided significance level set at 0.05.

## Results

### Baseline Characteristics (LVO)

**Table 1.**
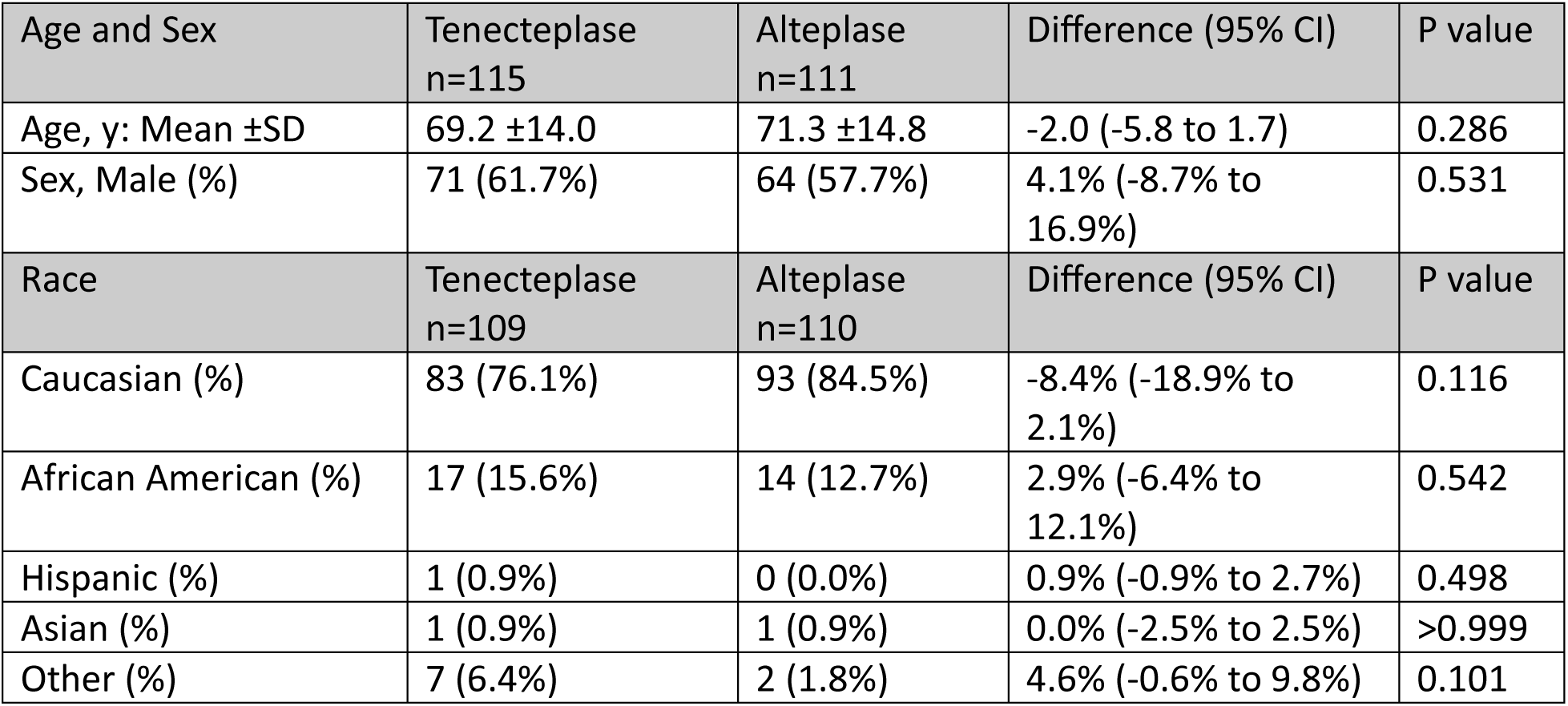
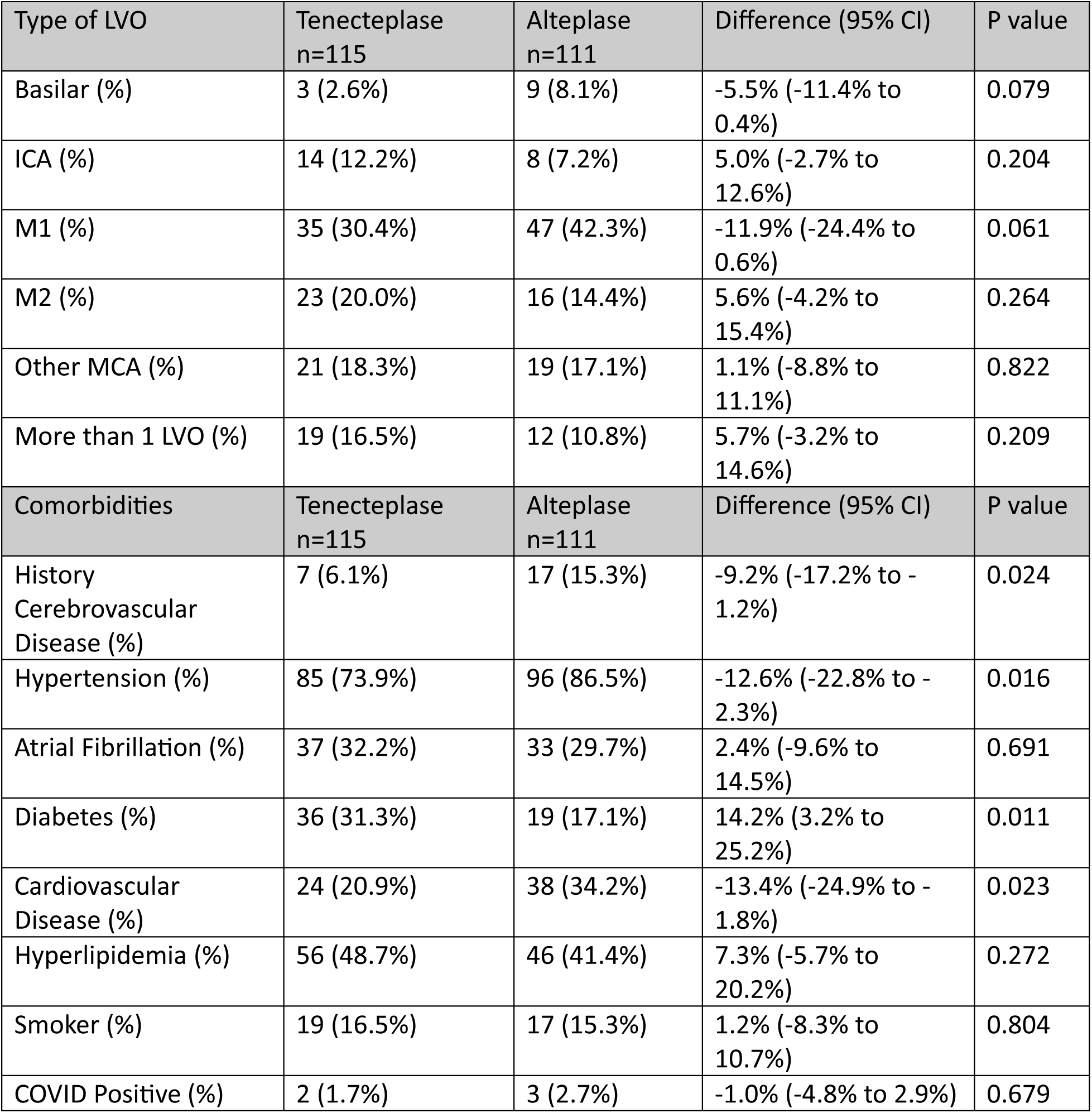
Baseline Characteristics Stratified by Thrombolytic Treatment (LVO Patients)

| Age and Sex | Tenecteplase<br>n=115 | Alteplase<br>n=111 | Difference (95% CI) | P value |
| --- | --- | --- | --- | --- |
| Age, y: Mean ±SD | 69.2 ±14.0 | 71.3 ±14.8 | -2.0 (-5.8 to 1.7) | 0.286 |
| Sex, Male (%) | 71 (61.7%) | 64 (57.7%) | 4.1% (-8.7% to 16.9%) | 0.531 |
| Race | Tenecteplase<br>n=109 | Alteplase<br>n=110 | Difference (95% CI) | P value |
| Caucasian (%) | 83 (76.1%) | 93 (84.5%) | -8.4% (-18.9% to 2.1%) | 0.116 |
| African American (%) | 17 (15.6%) | 14 (12.7%) | 2.9% (-6.4% to 12.1%) | 0.542 |
| Hispanic (%) | 1 (0.9%) | 0 (0.0%) | 0.9% (-0.9% to 2.7%) | 0.498 |
| Asian (%) | 1 (0.9%) | 1 (0.9%) | 0.0% (-2.5% to 2.5%) | >0.999 |
| Other (%) | 7 (6.4%) | 2 (1.8%) | 4.6% (-0.6% to 9.8%) | 0.101 |

| Type of LVO | Tenecteplase<br>n=115 | Alteplase<br>n=111 | Difference (95% CI) | P value |
| --- | --- | --- | --- | --- |
| Basilar (%) | 3 (2.6%) | 9 (8.1%) | -5.5% (-11.4% to 0.4%) | 0.079 |
| ICA (%) | 14 (12.2%) | 8 (7.2%) | 5.0% (-2.7% to 12.6%) | 0.204 |
| M1 (%) | 35 (30.4%) | 47 (42.3%) | -11.9% (-24.4% to 0.6%) | 0.061 |
| M2 (%) | 23 (20.0%) | 16 (14.4%) | 5.6% (-4.2% to 15.4%) | 0.264 |
| Other MCA (%) | 21 (18.3%) | 19 (17.1%) | 1.1% (-8.8% to 11.1%) | 0.822 |
| More than 1 LVO (%) | 19 (16.5%) | 12 (10.8%) | 5.7% (-3.2% to 14.6%) | 0.209 |
| Comorbidities | Tenecteplase<br>n=115 | Alteplase<br>n=111 | Difference (95% CI) | P value |
| History Cerebrovascular Disease (%) | 7 (6.1%) | 17 (15.3%) | -9.2% (-17.2% to -1.2%) | 0.024 |
| Hypertension (%) | 85 (73.9%) | 96 (86.5%) | -12.6% (-22.8% to -2.3%) | 0.016 |
| Atrial Fibrillation (%) | 37 (32.2%) | 33 (29.7%) | 2.4% (-9.6% to 14.5%) | 0.691 |
| Diabetes (%) | 36 (31.3%) | 19 (17.1%) | 14.2% (3.2% to 25.2%) | 0.011 |
| Cardiovascular Disease (%) | 24 (20.9%) | 38 (34.2%) | -13.4% (-24.9% to -1.8%) | 0.023 |
| Hyperlipidemia (%) | 56 (48.7%) | 46 (41.4%) | 7.3% (-5.7% to 20.2%) | 0.272 |
| Smoker (%) | 19 (16.5%) | 17 (15.3%) | 1.2% (-8.3% to 10.7%) | 0.804 |
| COVID Positive (%) | 2 (1.7%) | 3 (2.7%) | -1.0% (-4.8% to 2.9%) | 0.679 |

Patients who received TNK for AIS within 4.5 hours and large vessel occlusion had better 90-day functional outcomes and had higher incidence of reperfusion before thrombectomy compared to patients receiving alteplase. Patients receiving TNK (LVO and non-LVO) had higher incidence of being discharged home compared to patients that received alteplase.

The study included 476 AIS patients (TNK n=270; alteplase n=206), of whom 226 had LVO (TNK n=115; alteplase n=111) (Figure 1). Baseline demographics and initial stroke severity were similar between groups in LVO (median NIHSS 15 vs 15). Several comorbidities differed in LVO: prior cerebrovascular disease (6.1% TNK vs 15.3% alteplase), hypertension (73.9% vs 86.5%), diabetes (31.3% vs 17.1%), and cardiovascular disease (20.9% vs 34.2%). Occlusion patterns were broadly comparable, with M1 most common.

**Figure 1.**
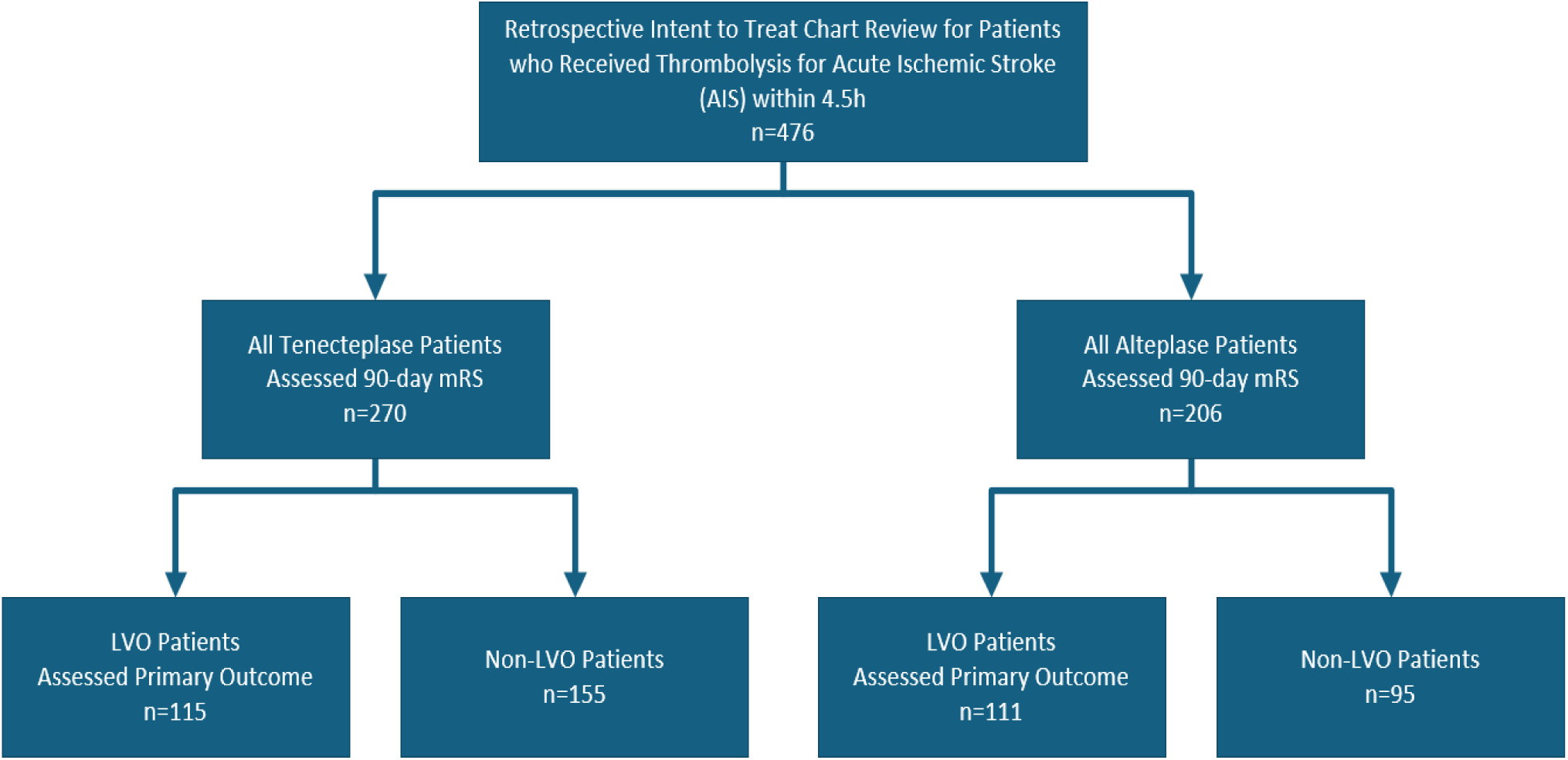
Cohort flow diagram showing sample-size breakdown (All AIS within 4.5h → Treatment group → LVO status).

### Primary Outcome (LVO)

**Table 2.**
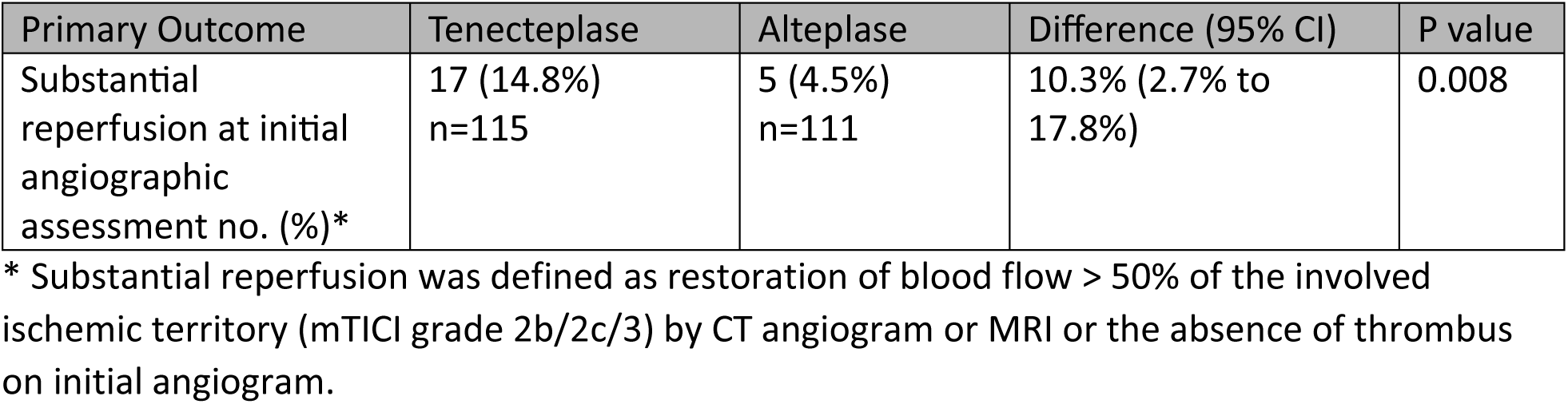
Primary Outcome (LVO Patients)

Early reperfusion prior to thrombectomy occurred in 17/115 (14.8%) TNK vs 5/111 (4.5%) alteplase (risk difference 10.3% [95% CI, 2.7–17.8], p=0.008). In multivariable binary logistic regression, TNK was associated with higher odds of early reperfusion compared with alteplase (adjusted OR 3.53, 95% CI 1.20–10.40, p=0.022). Figure 2 in the supplement is a forest plot showing the adjusted odds ratios with 95% confidence intervals based on binary logistic regression of the study’s predictors of early reperfusion prior to thrombectomy (LVO subgroup): treatment group (TNK vs alteplase), age, initial NIHSS, prior cerebrovascular disease (CVA/TIA), hypertension, atrial fibrillation, diabetes, cardiovascular disease, hyperlipidemia, and smoking status. A forest plot for a logistic regression model is a graphical representation used to visualize the odds ratios and their corresponding confidence intervals for the independent variables in the model.

**Figure 2.**
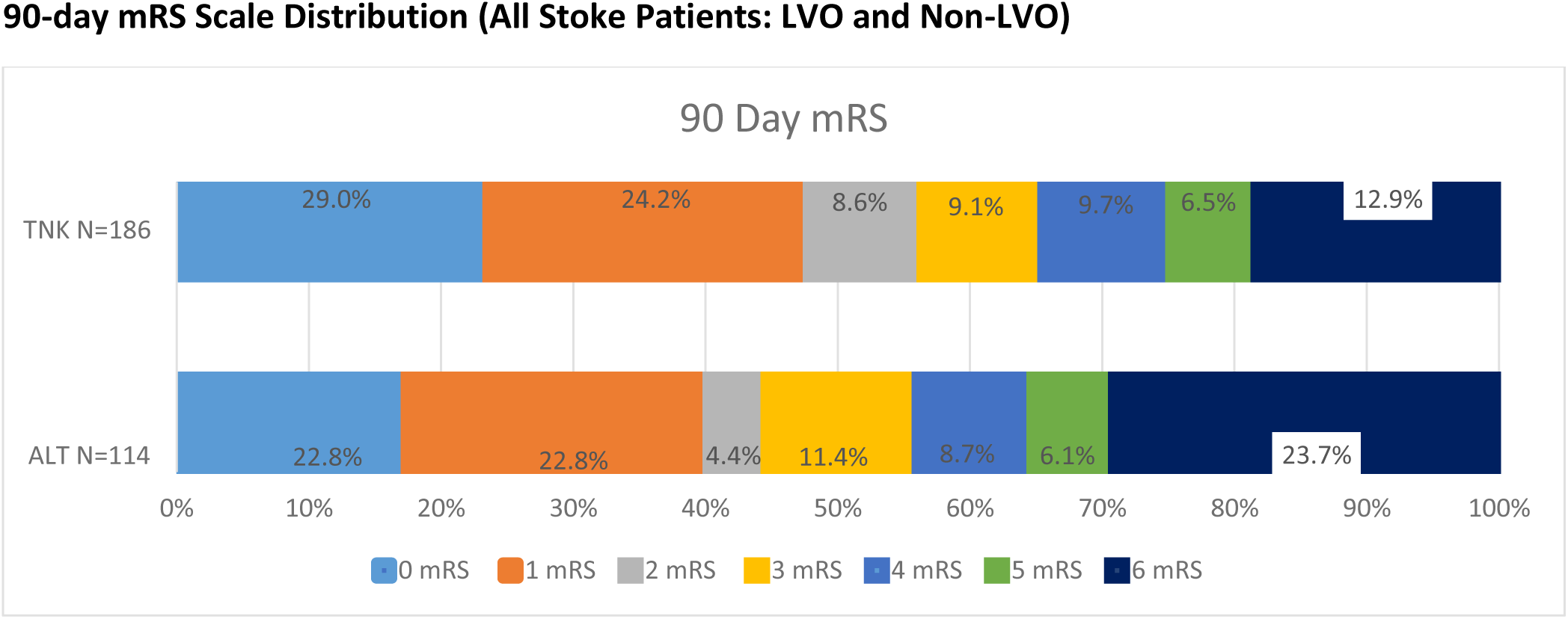
Modified Rankin Scale Scores at 90 days in the Intention to treat.

### Secondary and Safety Outcomes (LVO)

**Table 3.**
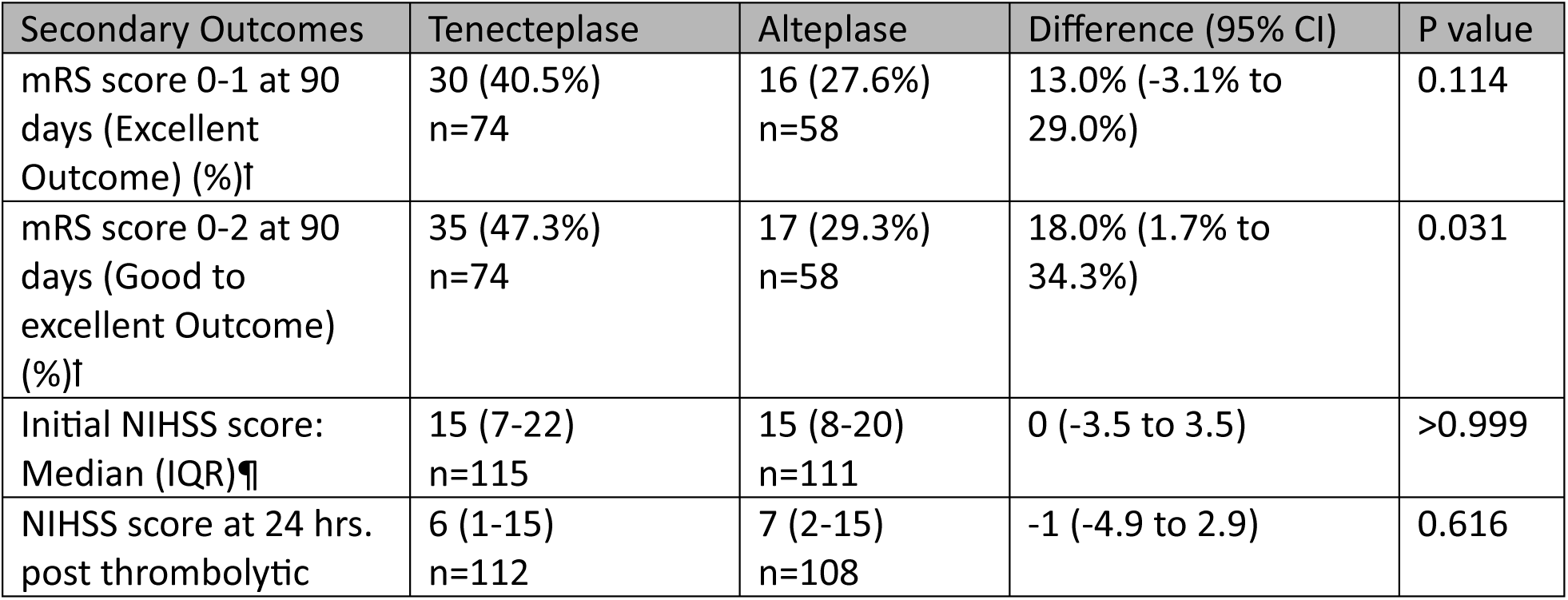
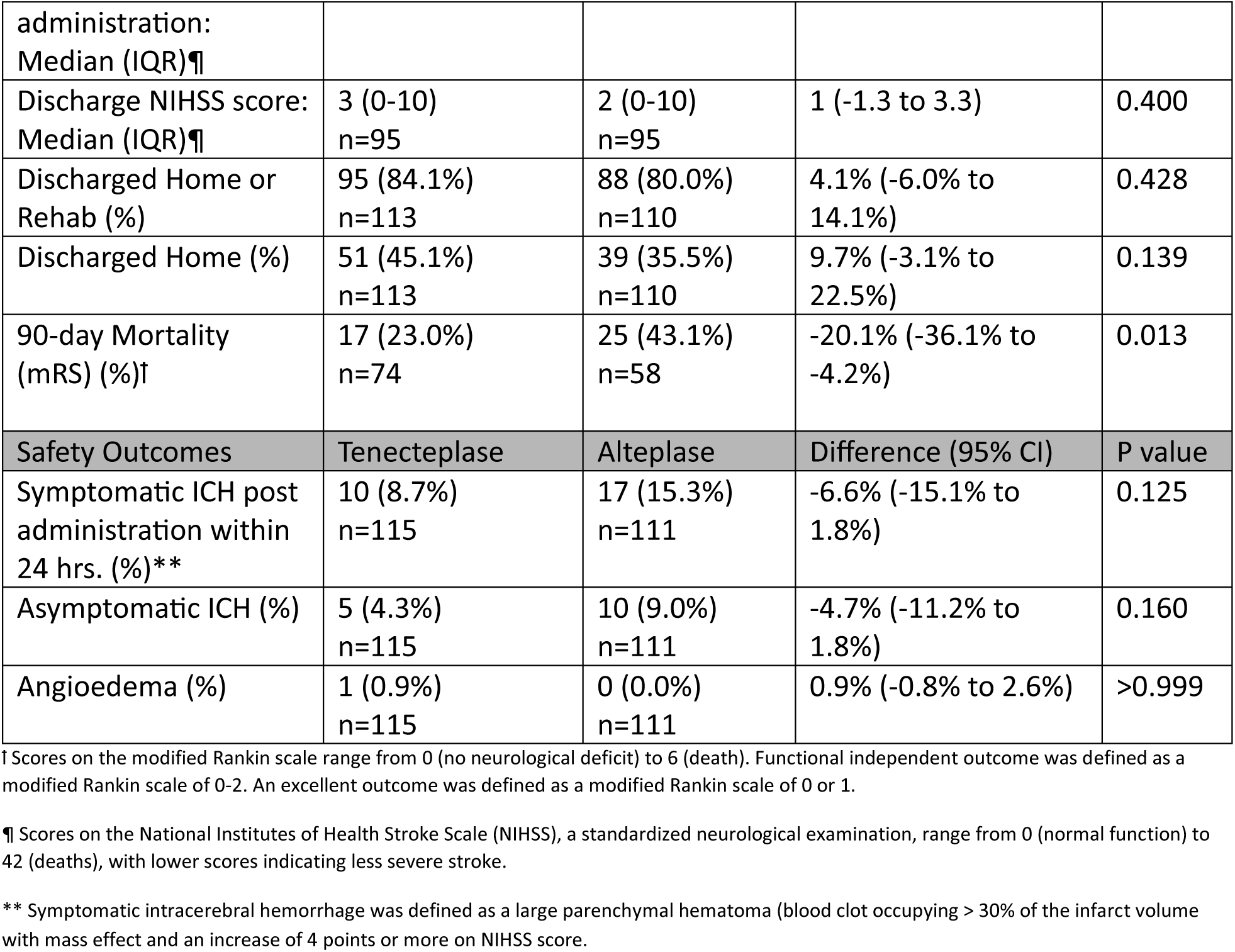
Secondary and Safety Outcomes (LVO Patients)

In LVO, 90-day good-to-excellent functional outcome (mRS 0–2) occurred in 47.3% with TNK vs 29.3% with alteplase (p=0.031); excellent outcome (mRS 0–1) was 40.5% vs 27.6% (p=0.114). In all AIS patients, excellent outcome occurred in 53.2% vs 45.6% (difference 7.6% [95% CI, −4.0 to 19.2], p=0.199), and good-to-excellent outcome in 61.8% vs 50.0% (difference 11.8% [95% CI, −0.3 to 23.4], p=0.044). Median 90-day mRS favored TNK (1 [IQR 0–4] vs 3 [1–5], p=0.046).

NIHSS score at 24 hours was lower with TNK (median 6 [IQR=1-15]) compared to alteplase (7 [2-15]) for the LVO group, but the difference was not statistically significant (p=0.616). Similarly, Discharge NIHSS score was higher with TNK (median 3 [IQR=0-10]) compared to alteplase (2 [0-10]) for the LVO group, but the difference was not statistically significant (p=0.400). While there was a higher percentage of patients discharged home with TNK compared to alteplase, 45.1% versus 35.5%, respectively, for the LVO group, there was not a statistically significant difference. When evaluating the 90-day mRS scale distribution between the two LVO groups, there was a trend to more patients in the TNK group having good to excellent outcome compared to the alteplase group, 47.3% versus 29.3%, respectively (p=0.031). Patients with excellent outcome at 90 days occurred in 40.5% with TNK compared to 27.6% with alteplase for the LVO group (p=0.114).

In LVO, 90-day mortality (mRS) was statistically lower with TNK compared to alteplase, 23.0% versus 43.1%, respectively, (difference -20.1% [95% CI, -36.1% to -4.2%], p=0.013). Also, in LVO, symptomatic ICH occurred in 8.7% with TNK vs 15.3% with alteplase (p=0.125); asymptomatic ICH 4.3% vs 9.0% (p=0.160). Across all AIS patients, combined ICH (symptomatic and asymptomatic) was less frequent with TNK (8.5% vs 15.0%, p=0.030), and mortality among symptomatic ICH at discharge was lower (2.2% vs 6.3%, p=0.033). Angioedema was uncommon (0.8% TNK vs 0%).

Symptomatic ICH occurred in 5.6% of all patients that received TNK and 9.2% in the alteplase group (difference -3.7% [95% CI, -8.5% to 1.1%], p=0.135). Asymptomatic ICH occurred in 3.0% in the TNK group compared to 5.8% in the alteplase group (difference -2.9% [95% CI, -6.6% to 0.9%], p=0.138) for all patients. There was not a statistical difference in bleeding complications between groups for LVO, however, there was a statistical difference between TNK and alteplase for the combined groups, 8.5% for TNK versus 15.0% for alteplase, (difference -6.5% [95% CI, -12.4% to -0.6%], p=0.030). In addition, when evaluating all patients that received thrombolytics, there was a statistical difference in symptomatic ICH mortality at discharge between the groups with 6 patients in the TNK group (2.2%) compared to 13 patients in the alteplase group (6.3%) (difference -4.1% [95% CI, -7.8% to -0.3%], p=0.033). Angioedema assessed for all patients after administration occurred in 2 patients in the TNK group (0.8%) and none in the alteplase group (0.0%) but was not statistically significant (difference 0.8% [95% CI, -0.3% to 1.8%], p=0.507). The 90-day mRS for mortality for all patients occurred in 12.9% in the TNK group and 23.7% in the alteplase group (difference -10.8% [95% CI, -20.0% to -1.60%], p=0.021).

### Demographics and Sample Characteristics (LVO and Non-LVO)

Across all treated patients (TNK, n=270; alteplase, n=206), baseline characteristics were broadly similar between groups (Table 4). Mean age did not differ significantly (68.2 ± 14.6 vs 70.0 ± 15.4 years; difference −1.8 years [95% CI, −4.5 to 1.0], p=0.210), nor did sex distribution (male: 53.7% vs 51.5%; difference 2.2% [95% CI, −6.8% to 11.3%], p=0.627). Initial stroke severity was comparable, with NIHSS score (median 8 [IQR 4–15] in the TNK group and 9 [IQR 5–16] in the alteplase group (difference −1 point [95% CI, −2.8 to 0.8], p=0.282).

**Table 4.** Demographics and Sample Characteristics (All Stroke Patients: LVO and Non-LVO)

| Demographics and Sample Characteristics | Tenecteplase<br>n=270 | Alteplase<br>n=206 | Difference (95% CI) | P value |
| --- | --- | --- | --- | --- |
| Age, y: Mean ±SD | 68.2 ±14.6 | 70.0 ±15.4 | -1.8 (-4.5 to 1.0) | 0.210 |
| Sex, Male (%) | 145 (53.7%) | 106 (51.5%) | 2.2% (-6.8% to 11.3%) | 0.627 |
| Initial NIHSS score: Median (IQR)* | 8 (4-15) | 9 (5-16) | -1 (-2.8 to 0.8) | 0.282 |
| Race | Tenecteplase<br>n=263 | Alteplase<br>n=204 | Difference (95% CI) | P value |
| Caucasian (%) | 212 (80.6%) | 174 (85.3%) | -4.7% (-11.5% to 2.1%) | 0.178 |
| African American (%) | 33 (12.5%) | 23 (11.3%) | 1.3% (-4.6% to 7.2%) | 0.673 |
| Hispanic (%) | 3 (1.1%) | 1 (0.5%) | 0.7% (-1.0% to 2.2%) | 0.635 |
| Asian (%) | 3 (1.1%) | 2 (1.0%) | 0.2% (-1.7% to 2.0%) | >0.999 |
| Other (%) | 11 (4.2%) | 4 (2.0%) | 2.2% (-0.9% to 5.3%) | 0.198 |

| Comorbidities | Tenecteplase<br>n=253 | Alteplase<br>n=202 | Difference (95% CI) | P value |
| --- | --- | --- | --- | --- |
| History Cerebrovascular Disease (%) | 23 (9.1%) | 26 (12.9%) | -3.8% (-9.6% to 2.0%) | 0.203 |
| Hypertension (%) | 204 (80.6%) | 165 (81.7%) | -1.1% (-8.3% to 6.2%) | 0.776 |
| Atrial Fibrillation (%) | 50 (19.8%) | 49 (24.3%) | -4.5% (-12.2% to 3.2%) | 0.252 |
| Diabetes (%) | 77 (30.4%) | 43 (21.3%) | 9.1% (1.1% to 17.1%) | 0.025 |
| Cardiovascular Disease (%) | 52 (20.6%) | 68 (33.7%) | -13.1% (-21.3% to -4.9%) | 0.002 |
| Hyperlipidemia (%) | 123 (48.6%) | 89 (44.1%) | 4.6% (-4.7% to 13.8%) | 0.332 |
| Smoker (%) | 58 (22.9%) | 34 (16.8%) | 6.1% (-1.2% to 13.4%) | 0.102 |
| COVID (%) | 9 (3.6%) | 4 (2.0%) | 1.6% (-1.4% to 4.6%) | 0.402 |

\*Scores on the National Institutes of Health Stroke Scale (NIHSS), a standardized neurological examination, range from 0 (normal function) to 42 (deaths), with lower scores indicating less severe stroke.

Racial composition was similar, with no statistically significant differences for Caucasian (80.6% vs 85.3%; difference −4.7% [95% CI, −11.5% to 2.1%], p=0.178), African American (12.5% vs 11.3%; difference 1.3% [95% CI, −4.6% to 7.2%], p=0.673), Hispanic (1.1% vs 0.5%; difference 0.7% [95% CI, −1.0% to 2.2%], p=0.635), Asian (1.1% vs 1.0%; difference 0.2% [95% CI, −1.7% to 2.0%], p>0.999), or Other (4.2% vs 2.0%; difference 2.2% [95% CI, −0.9% to 5.3%], p=0.198).

Two comorbidities differed significantly between groups. Diabetes was more prevalent among patients receiving TNK (30.4% vs 21.3%; difference 9.1% [95% CI, 1.1% to 17.1%], p=0.025). Conversely, cardiovascular disease was less common with TNK (20.6% vs 33.7%; difference −13.1% [95% CI, −21.3% to −4.9%], p=0.002). There were no significant differences for prior cerebrovascular disease (9.1% vs 12.9%; difference −3.8% [95% CI, −9.6% to 2.0%], p=0.203), hypertension (80.6% vs 81.7%; difference −1.1% [95% CI, −8.3% to 6.2%]; p=0.776), atrial fibrillation (19.8% vs 24.3%; difference −4.5% [95% CI, −12.2% to 3.2%], p=0.252), hyperlipidemia (48.6% vs 44.1%; difference 4.6% [95% CI: −4.7% to 13.8%], p=0.332), smoking (22.9% vs 16.8%; difference 6.1% [95% CI, −1.2% to 13.4%], p=0.102), or COVID status (3.6% vs 2.0%; difference 1.6% [95% CI, −1.4% to 4.6%]; p=0.402). Subgroup denominators vary slightly due to missing data, as reflected in the table.

### Secondary and Safety Outcomes (LVO and Non-LVO)

**Table 5.** Secondary and Safety Outcomes (All Stroke Patients: LVO and Non-LVO)

| Secondary Outcomes | Tenecteplase | Alteplase | Difference (95% CI) | P value |
| --- | --- | --- | --- | --- |
| Door-to-Needle Time, mins: Median (IQR) | 34 (27-44)<br>n=178 | 45 (36-53)<br>n=110 | -11 (-14.4 to -7.6) | <0.001 |
| 90-day mRS: Median (IQR) | 1 (0-4)<br>n=186 | 3 (1-5)<br>n=114 | 0 (-1 to 0) | 0.046 |
| mRS score 0-1 at 90 days (Excellent Outcome) (%)† | 99 (53.2%)<br>n=186 | 52 (45.6%)<br>n=114 | 7.6% (-4.0% to 19.2%) | 0.199 |
| mRS score 0-2 at 90 days (Good to Excellent Outcome) (%)† | 115 (61.8%)<br>n=186 | 57 (50.0%)<br>n=114 | 11.8% (-0.3% to 23.4%) | 0.044 |
| mRS score 6 at 90 days (%) | 24 (12.9%)<br>n=186 | 27 (23.7%)<br>n=114 | -10.8% (-20.0% to -1.6%) | 0.021 |
| Discharged Home or Rehab (%) | 241 (89.9%)<br>n=268 | 178 (86.8%)<br>n=205 | 3.1% (-2.8% to 9.0%) | 0.301 |
| Discharged Home (%) | 156 (58.2%)<br>n=268 | 100 (48.8%)<br>n=205 | 9.4% (0.4% to 18.5%) | 0.041 |
| 90-day Mortality (mRS) (%)† | 24 (12.9%)<br>n=186 | 27 (23.7%)<br>n=114 | -10.8% (-20.0% to -1.60%) | 0.021 |
| Safety Outcomes | Tenecteplase | Alteplase | Difference (95% CI) | P value |
| Symptomatic ICH post administration within 24 hrs. (%)** | 15 (5.6%)<br>n=270 | 19 (9.2%)<br>n=206 | -3.7% (-8.5% to 1.1%) | 0.135 |
| Asymptomatic ICH post administration within 24hrs. (%) | 8 (3.0%)<br>n=270 | 12 (5.8%)<br>n=206 | -2.9% (-6.6 to 0.9%) | 0.138 |
| Combined ICH post administration 24 hrs. (%) | 23 (8.5%)<br>n=270 | 31 (15.0%)<br>n=206 | -6.5% (-12.4% to -0.6%) | 0.030 |
| Mortality Symptomatic ICH discharge (%)** | 6 (2.2%)<br>n=270 | 13 (6.3%)<br>n=206 | -4.1% (-7.8 to -0.3%) | 0.033 |
| Angioedema (%) | 2 (0.8%)<br>n=266 | 0 (0.0%)<br>n=205 | 0.8% (-0.3% to 1.8%) | 0.507 |

† Scores on the modified Rankin scale range from 0 (no neurological deficit) to 6 (death). Functional independent outcome was defined as a modified Rankin scale of 0-2. An excellent outcome was defined as a modified Rankin scale of 0 or 1.

Among all AIS patients, DTN was shorter with TNK (median 34 [IQR 27–44] minutes) than with alteplase (45 [36–53] minutes), a −11-minute difference (95% CI, −14.4 to −7.6; p<0.001). Evaluating the scale distribution of 90-day mRS in all patients that received thrombolytics showed patients with excellent outcome (mRS 0-1) occurred in 53.2% of all patients that received TNK versus 45.6% for the alteplase group (difference 7.6% [95% CI, -4.0% to 19.2%], p=0.199). Good to excellent outcome (mRS 0-2) occurred in 61.8% in all patients that received TNK and 50.0% in the alteplase group (statistically significant difference 11.8% [95% CI, -0.3% to 23.4%], p=0.044). Figure 1 in the supplement shows the Modified Rankin Scale Scores at 90 days with the Intention to treat as shown in the scale distribution of the modified Rankin scale scores at 90 days. There was a statistical difference in all the patients that were discharged home with 58.2% in the TNK group versus 48.8% in the alteplase group (difference 9.4% [95% CI, 0.4% to 18.5%], p=0.041).

### Adjusted Associations for 90-Day Excellent Outcome (LVO Subgroup)

In the LVO subgroup logistic model for excellent outcome (mRS 0–1), treatment group was not a significant independent predictor (TNK vs alteplase: OR 1.26 [95% CI, 0.52–3.05], p=0.601). Lower initial stroke severity and younger age were associated with higher odds of excellent outcome (initial NIHSS per point: OR 0.91 [95% CI, 0.86–0.97], p=0.002; age per 10 years: OR 0.70 [95% CI, 0.50–0.96], p=0.027). Hyperlipidemia was positively associated (OR 2.50 [95% CI, 1.05–5.93], p=0.038). Figure 3 in the Supplement is a forest plot showing the adjusted odds ratios with 95% confidence intervals based on binary logistic regression of the study’s predictors of 90-day excellent outcome: treatment group (TNK vs alteplase), age, initial NIHSS, prior cerebrovascular disease (CVA/TIA), hypertension, atrial fibrillation, diabetes, cardiovascular disease, hyperlipidemia, and smoking status.

### Adjusted Associations for 90-Day Good Outcome (LVO Subgroup)

In the LVO subgroup logistic model for good outcome (mRS 0–2), treatment group was not a significant independent predictor (TNK vs alteplase: OR 1.69 [95% CI, 0.71–4.00], p=0.236). Lower initial stroke severity and younger age were associated with higher odds of good outcome (initial NIHSS per point: OR 0.91 [95% CI, 0.85–0.96], p=0.001; age per 10 years: OR 0.73 [95% CI, 0.53–1.00], p=0.048). Hyperlipidemia was positively associated (OR 2.55 [95% CI, 1.08–6.03], p=0.033). Figure 4 in the Supplement is a forest plot showing the adjusted odds ratios with 95% confidence intervals based on binary logistic regression of the study’s predictors of 90-day good outcome: treatment group (TNK vs alteplase), age, initial NIHSS, prior cerebrovascular disease (CVA/TIA), hypertension, atrial fibrillation, diabetes, cardiovascular disease, hyperlipidemia, and smoking status.

### Adjusted Associations for 90-Day Ordinal Outcome (LVO Subgroup)

In the LVO subgroup logistic model for ordinal outcome mRS, treatment group was not a significant independent predictor (TNK vs alteplase: OR 0.58 [95% CI, 0.29–1.17], p=0.129). Higher initial stroke severity and older age were associated with higher odds of higher 90-day mRS ordinal outcomes (initial NIHSS per point: OR 1.09 [95% CI, 1.05–1.14], p<0.001; age per 10 years: OR 1.43 [95% CI, 1.12–1.82], p=0.004). Hyperlipidemia was negatively associated with higher odds of higher 90-day mRS ordinal outcomes (OR 0.44 [95% CI, 1.08–6.03], p=0.033). Figure 5 in the Supplement is a forest plot showing the adjusted odds ratios with 95% confidence intervals based on ordinal logistic regression of the study’s predictors of 90-day mRS ordinal outcome: treatment group (TNK vs alteplase), age, initial NIHSS, prior cerebrovascular disease (CVA/TIA), hypertension, atrial fibrillation, diabetes, cardiovascular disease, hyperlipidemia, and smoking status.

### Adjusted Associations for Mortality Related to a Bleed Outcome (LVO Subgroup)

In the LVO subgroup logistic model for mortality related to a bleed, treatment group was not a significant independent predictor (TNK vs alteplase: OR 0.25 [95% CI, 0.04–1.52], p=0.132). Hyperlipidemia was negatively associated with higher odds of mortality related to a bleed (OR 0.14 [95% CI, 0.03–0.76], p=0.023). Figure 6 in the Supplement is a forest plot showing the adjusted odds ratios with 95% confidence intervals based on binary logistic regression of the study’s predictors of mortality related to a bleed: treatment group (TNK vs alteplase), age, initial NIHSS, prior cerebrovascular disease (CVA/TIA), hypertension, atrial fibrillation, diabetes, cardiovascular disease, hyperlipidemia, and smoking status.

### Adjusted Associations for All-Cause Mortality 90-Day mRS 6 (LVO Subgroup)

In the LVO subgroup logistic model for 90-day all-cause mortality (mRS 6), treatment group was not a significant independent predictor (TNK vs alteplase: OR 0.38 [95% CI, 0.14–1.05], p=0.061). Higher initial stroke severity and older age were associated with higher odds of 90-day all-cause mortality (initial NIHSS per point: OR 1.12 [95% CI, 1.06–1.19], p<0.001; age per 10 years: OR 1.70 [95% CI, 1.19–2.41], p=0.003). Figure 7 in the Supplement is a forest plot showing the adjusted odds ratios with 95% confidence intervals based on binary logistic regression of the study’s predictors of 90-day all-cause mortality: treatment group (TNK vs alteplase), age, initial NIHSS, prior cerebrovascular disease (CVA/TIA), hypertension, atrial fibrillation, diabetes, cardiovascular disease, hyperlipidemia, and smoking status.

### Predictive Margins (Marginal Plots)

Marginal plots are graphical representations of the marginal effects calculated from regression models. They help visualize how changes in independent variables affect the predicted values of the dependent variable while holding other variables constant. This is particularly useful for interpreting complex models with multiple predictors.

#### Early Reperfusion Prior to Thrombectomy (LVO Subgroup)

TNK was independently associated with higher odds of early reperfusion (OR 3.53 [95% CI, 1.20–10.40], p=0.022). Age (per 10 years; OR 1.04 [95% CI, 0.75–1.46], p=0.797) and initial NIHSS (per point; OR 0.99 [95% CI, 0.93–1.05], p=0.721) were not significant predictors. Other covariates had wide CIs that crossed 1.00 (e.g., CVA/TIA OR 0.54 [95% CI, 0.064–4.59], p=0.574; hypertension OR 0.46 [95% CI, 0.16–1.29], p=0.140).

Predicted probabilities of early reperfusion calculated by the model were higher with TNK than with alteplase across the displayed ranges of age and baseline NIHSS. The absolute gap between agents was relatively consistent across age strata and NIHSS bands, mirroring the adjusted odds ratio advantage for TNK referenced above. Figures 8–9 in the Appendix are marginal plots provided to show the predicted probabilities at various levels of the model predictors.

#### 90-Day mRS (Binary Outcomes) (LVO Subgroup)

Treatment with TNK was not a significant independent predictor of excellent outcome (OR 1.26 [95% CI, 0.52–3.05], p=0.601). Younger age (per 10 years; OR 0.70 [95% CI, 0.50–0.96], p=0.027) and lower initial NIHSS (per point; OR 0.91 [95% CI, 0.86–0.97], p=0.002) were associated with increased odds of achieving mRS 0–1. Hyperlipidemia was also positively associated (OR 2.50 [95% CI, 1.05–5.93], p=0.038).

TNK was not a significant predictor of mRS 0–2 (OR 1.69 [95% CI, 0.71–4.00], p=0.236). Lower initial NIHSS remained strongly associated with better outcomes (OR 0.91 [95% CI, 0.85–0.96], p=0.001). Age showed a borderline association suggesting lower odds of mRS 0–2 with increasing age (OR 0.73 [95% CI, 0.53–1.00], p=0.048). Hyperlipidemia again favored better outcomes (OR 2.55 [95% CI, 1.08–6.03], p=0.033).

For excellent outcome (mRS 0–1) and good outcome (mRS 0–2), margins decreased monotonically with increasing age and higher initial NIHSS. Across comparable ages and NIHSS values, TNK and alteplase yielded broadly similar predicted probabilities, consistent with non-significant adjusted treatment odds ratios. The between-group margins occasionally favored TNK at lower NIHSS and younger ages, but 95% CIs overlapped throughout most of the range. Figures 10–13 in the Appendix are marginal plots provided to show the predicted probabilities at various levels of the model predictors.

#### 90-Day mRS (Ordinal Outcome) (LVO Subgroup)

In the proportional-odds ordinal logistic model (higher OR implies a shift toward worse mRS), TNK was not significantly associated with the distribution of mRS (OR 0.58 [95% CI, 0.29–1.17], p=0.129). Age (per 10 years) and initial NIHSS (per point) were associated with worse functional status (age OR 1.43 [95% CI, 1.12–1.82], p=0.004; NIHSS OR 1.09 [95% CI, 1.05–1.14], p<0.001). Hyperlipidemia was associated with a shift toward better outcomes (OR 0.44 [95% CI, 0.23– 0.86], p=0.017).

#### Mortality Outcomes (LVO Subgroup)

For mortality related to bleeding, events were infrequent, and CIs were wide. TNK showed a nonsignificant trend toward lower odds of mortality related to a bleed (OR 0.25 [95% CI, 0.04– 1.52], p=0.132). Hyperlipidemia was associated with lower odds (OR 0.14 [95% CI, 0.03–0.76], p=0.023). Initial NIHSS showed a positive but nonsignificant association (OR 1.09 [95% CI, 0.99– 1.20], p=0.083). Figure 14 in the supplement is a marginal plot provided to show the predicted probabilities by age at various levels of the model predictors.

TNK was associated with a lower—though not statistically significant—odds of death for 90 days (mRS 6) (OR 0.38 [95% CI, 0.14–1.05], p=0.061). Older age and higher stroke severity were significantly associated with mortality (age per 10 years OR 1.70 [95% CI, 1.19–2.41], p=0.003; NIHSS per point OR 1.12 [95% CI, 1.06–1.19], p<0.001). Hyperlipidemia showed a protective trend though not statistically significant in this case (OR 0.39 [95% CI, 0.15–1.04], p=0.059).

Predicted probabilities of death by 90 days (mRS 6) increased with age and baseline NIHSS. TNK exhibited numerically lower predicted mortality across a range of ages and severities, concordant with the adjusted OR below 1; however, uncertainty bands were wide, particularly at extreme age and NIHSS values. Figure 15 in the supplement is a marginal plot provided to show the predicted probabilities by initial NIHSS at various levels of the model predictors.

#### Overall Interpretation

Predictive margins recast the regression findings into patient-centered probabilities: TNK confers a consistent advantage for early reperfusion in LVO, while downstream functional outcomes are chiefly determined by age and initial stroke severity. These visuals can help clinicians appreciate how absolute risks change across age and NIHSS while reinforcing that the mechanistic benefit (early reperfusion) is not always sufficient to overcome the prognostic weight of baseline severity on 90-day outcomes.

## Discussion

In a large integrated health system, TNK 0.25 mg/kg was associated with significantly higher early reperfusion prior to thrombectomy among LVO patients and with shorter DTN across all AIS patients, without excess bleeding. Functional outcomes favored TNK in several analyses (including LVO mRS 0–2 and median 90-day mRS), while adjusted odds of excellent outcome did not differ significantly between agents after accounting for key covariates (age, stroke severity, and comorbidities). Findings align with randomized evidence supporting TNK as an effective alternative to alteplase and suggest potential workflow advantages related to single-bolus administration.

Also, across additional multivariable models, TNK consistently improved the proximal, mechanistic target (early reperfusion before thrombectomy) while not demonstrating independent superiority on 90-day mRS after adjusting for age and initial NIHSS. Age and baseline stroke severity were the most robust determinants of functional outcomes and mortality. The repeated association between hyperlipidemia and favorable outcomes likely reflects confounding by statin exposure or overall cardiovascular risk management rather than a causal protective effect; nevertheless, the signal was directionally consistent across models. Taken together with shorter door-to-needle times and comparable safety, these data support the clinical and operational advantages of TNK in real-world community hospital LVO and AIS care.

## Conclusion

Tenecteplase 0.25 mg/kg is an effective and practical alternative to alteplase for AIS. In real-world community practice it improved pre-thrombectomy reperfusion in LVO, shortened door-to-needle times, and demonstrated comparable or better safety without increasing hemorrhagic complications.

This retrospective observational study comparing TNK to alteplase for AIS reinforces the current published evidence demonstrating TNK dosed at 0.25 mg/kg IV with max 25 mg IV is safe and effective alternative to alteplase AIS standard dosing. The primary outcome in this observational study did show similar results to the EXTEND-IA study with TNK resulting in a 10.3% reperfusion rate difference compared to Alteplase for LVO prior to thrombectomy which was statistically significant p=0.008. The number needed to treat (NNT) to see 50% reperfusion prior to thrombectomy in the TNK group was 7 in this study. This was slightly higher than the EXTEND IA trial which was 5. Also, in the adjusted logistic regression model, the odds of achieving reperfusion prior to thrombectomy were 3.53 times higher with tenecteplase than with alteplase (OR = 3.53 [95% CI, 1.20 to 10.40], p=0.022), resulting in an absolute improvement of approximately 10 percentage points (model-based average marginal effect of 9.7 percentage points) which closely corresponds to the observed 10.3% absolute difference. These findings are consistent with EXTEND-IA and reinforce that 0.25 mg/kg IV (max 25 mg) is a safe, efficacious dose.

This study also showed TNK has similar safety outcomes to alteplase for AIS. Operationally, tenecteplase reduced door-to-needle time by 11 minutes (p < 0.001), which is clinically meaningful given established time-outcome relationships and may have contributed to favorable secondary outcomes (e.g., higher 90-day mRS 0-2 in LVO). This could be due to the ease of administration of TNK compared to alteplase. The AcT trial sub-analysis showed that for every 30-minute reduction in onset to needle time, an additional 2% achieved an excellent outcome. For every 60 min reduction in door-to-needle time, an additional 1% achieved an excellent outcome.^23^ The ease of administration allows for smoother transition during a transfer to a comprehensive center for thrombectomy. Although this retrospective design and missing 90-day mRS data introduce limitations, the study reflects diverse, real-world community hospital utilization with safety similar to alteplase.

As of March 3^rd^ 2025 the FDA approved tenecteplase (TNKase®) for the treatment of AIS.^24^ The recommended package insert dosing was based off the AcT study dosing algorithm which utilizes weight ranges rounding to the nearest 2.5 mg.^20^ Patient’s less than 60 kg would receive 15 mg on the low end and there is a maximum dose of 25 mg on the high end. The 0.25 mg/kg TNK IV dose was not included in the package insert and is the dose currently recommended in the 2022 European stroke guidelines. The AcT trial did not include or report safety and efficacy outcomes specific to this dosing algorithm which is referenced in the appendix of the AcT study.^20^ The AcT study conclusion was 0.25 mg/kg IV TNK is a reasonable alternative to alteplase for all patients presenting with acute ischemic stroke who meet standard criteria for thrombolysis.^20^ The NOR-TEST-2 trial evaluated 0.4 mg/kg IV TNK and was terminated early based on overall ICH bleeds at 21%.^26^ Based on the approved dosing label, extremely low weight patients could approach the NOR-TEST-2 study dose based on the fixed 15 mg TNK dose The AcT trial did not evaluate each specific dose range included in the appendix dosing chart. This raises the questions, did the AcT study include enough patients in the < 60 kg category to validate the safety of the 15 mg dose in extremely low weight patients included in the package insert dosing for TNK. This study provides additional evidence to support 0.25 mg/kg IV TNK dose. The dosing protocol used in this trial was based on actual body weight dosed at 0.25 mg/kg IV with max of 25 mg rounded to the nearest 1 mg. See appendix 1. The ATTEST-2 study also used a weight-based dosing at 0.25 mg/kg IV max 25mg rounding to the nearest 0.5 mg. Our study had a limited number of patients in the low weight category and further studies may be needed to evaluate the safety in this population. The data from this study, coupled with easier administration and faster treatment times, further supports TNK as a preferred thrombolytic for eligible AIS patients.

Shown is the scale distribution of the modified Rankin scale scores at 90 days. Scores range from 0 to 6, with 0 indicating no neurological deficit. 1 no clinical significant disability, 2 slight disability (able to handle own affairs without assistance but unable to carry out all previous activities), 3 moderate disability requiring some help ( e.g. with shopping, cleaning, and finances but able to walk unassisted), 4 moderately severe disability (unable to attend to bodily needs without assistance and unable to walk unassisted), 5 severe disability (requiring constant nursing care and attention), and 6 death. Patients in the TNK group had a median score of 1, as compared with a median score of 3 among patients in the Alteplase group (95% CI: -1 to 0, p=0.046).

## Data Availability

Data is available

## Acknowledgments

The authors would like to acknowledge David Ball, Amanda Belt, Joanna Caranante, Stephanie Conners, Charlie Guastella, Ann Lumia, Laura Moore, Saurabh Narkhede, Haylee Regan for their generous support with this project and Tracy Johns as a reviewer for this paper.

## Supplement

**Figure 3.**
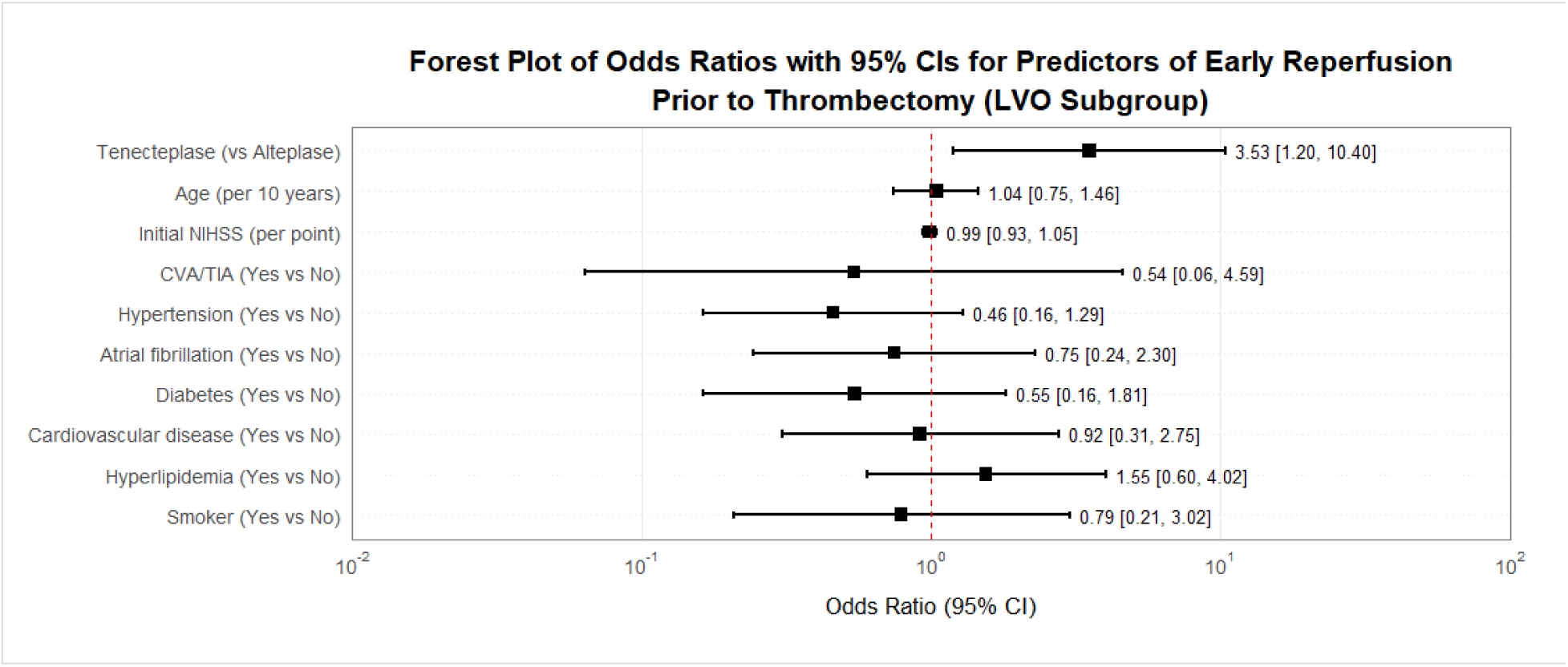
Adjusted odds ratios for predictors of early reperfusion prior to thrombectomy (LVO subgroup) based on binary logistic regression.

**Figure 4.**
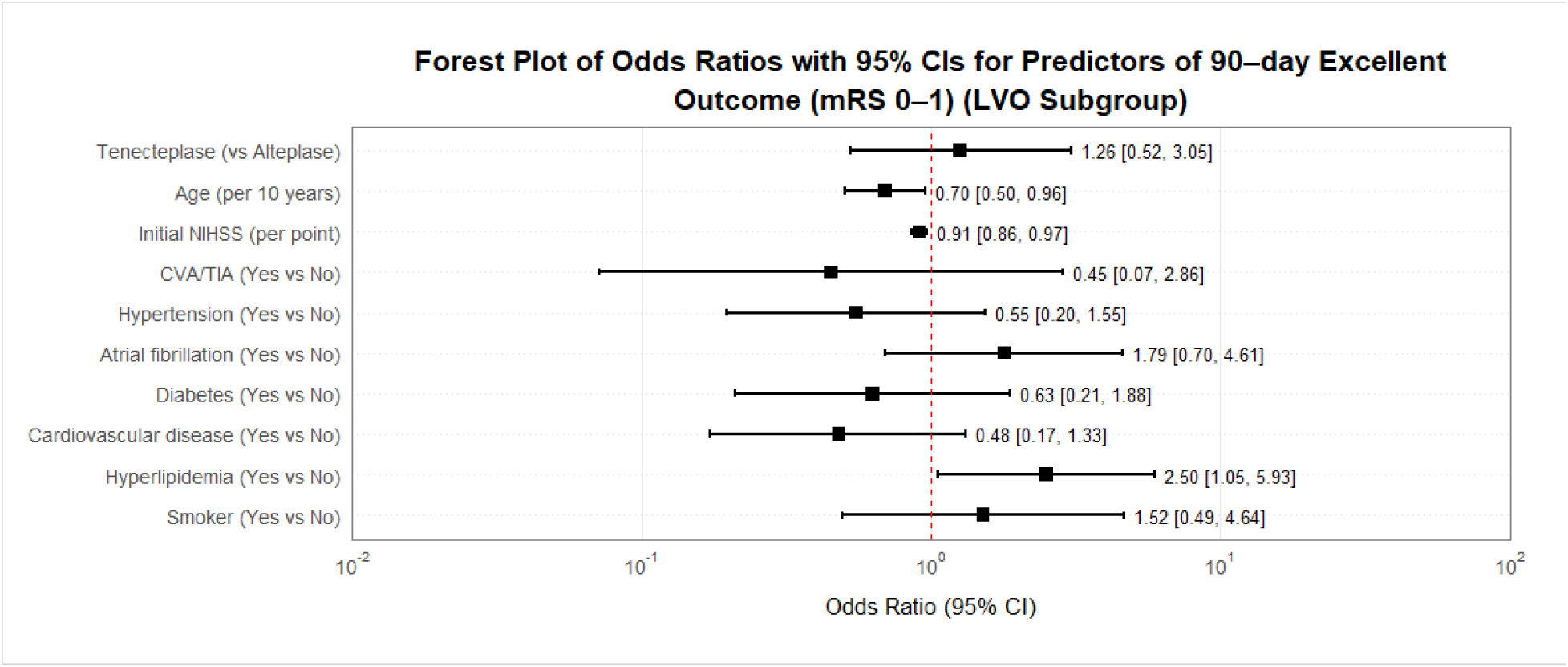
Adjusted odds ratios for predictors of 90-day excellent outcome (mRS 0–1) (LVO subgroup) based on binary logistic regression.

**Figure 5.**
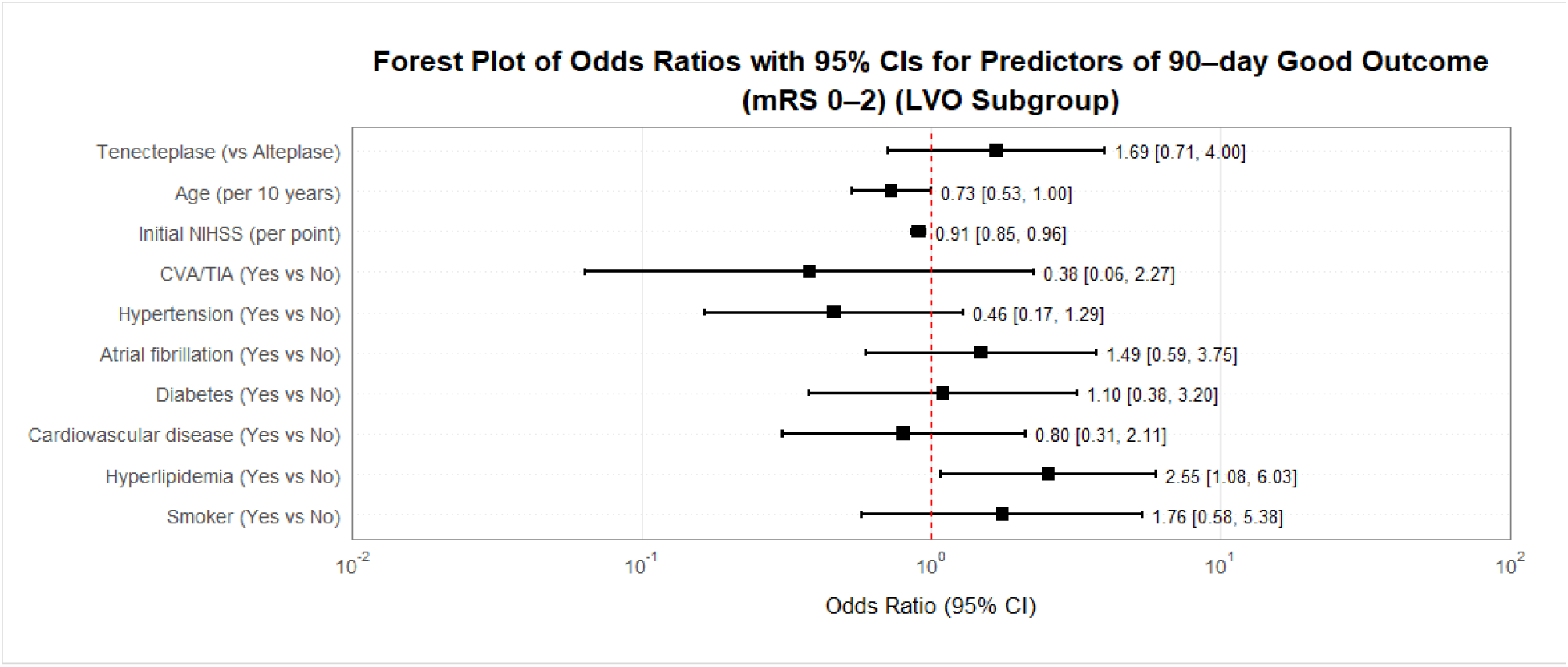
Adjusted odds ratios for predictors of 90-day good outcome (mRS 0–2) (LVO subgroup) based on binary logistic regression.

**Figure 6.**
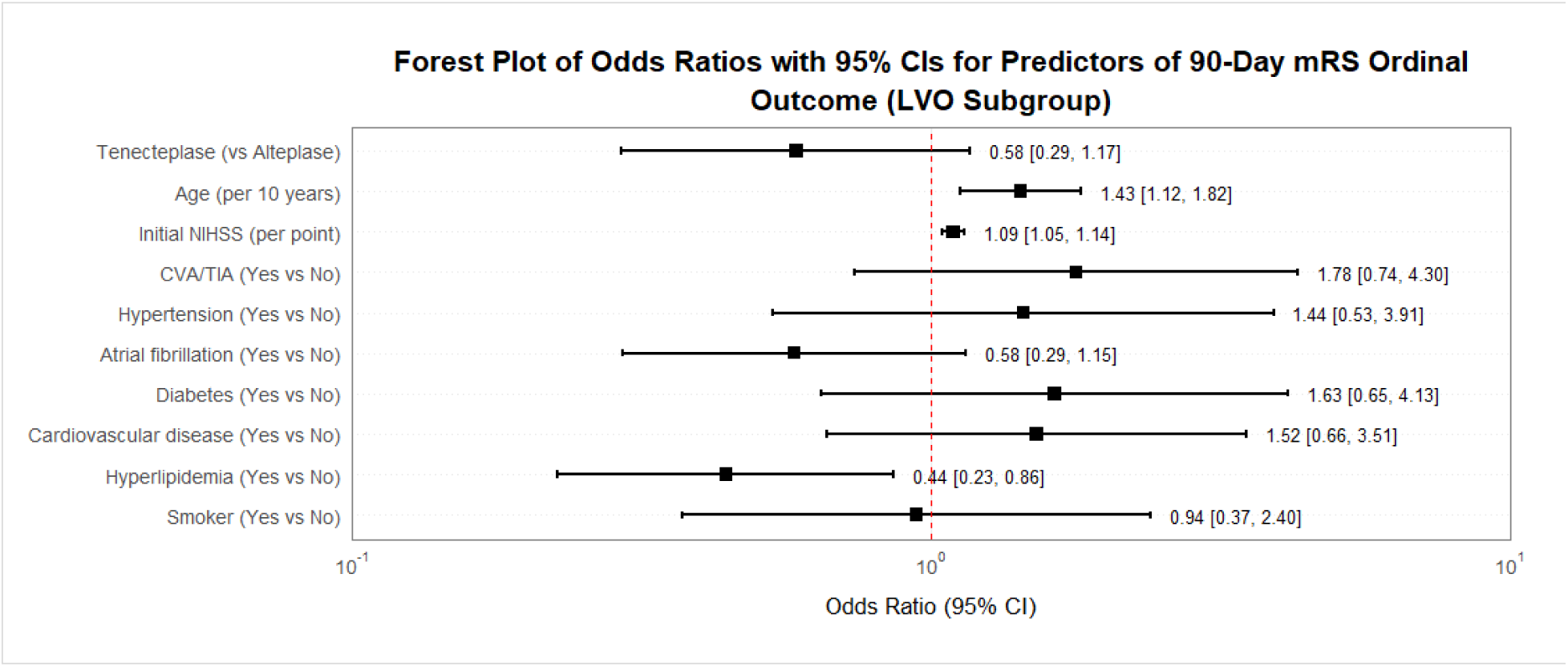
Adjusted odds ratios for predictors of 90-day mRS ordinal outcome (LVO subgroup) based on proportional-odds ordinal logistic regression.

**Figure 7.**
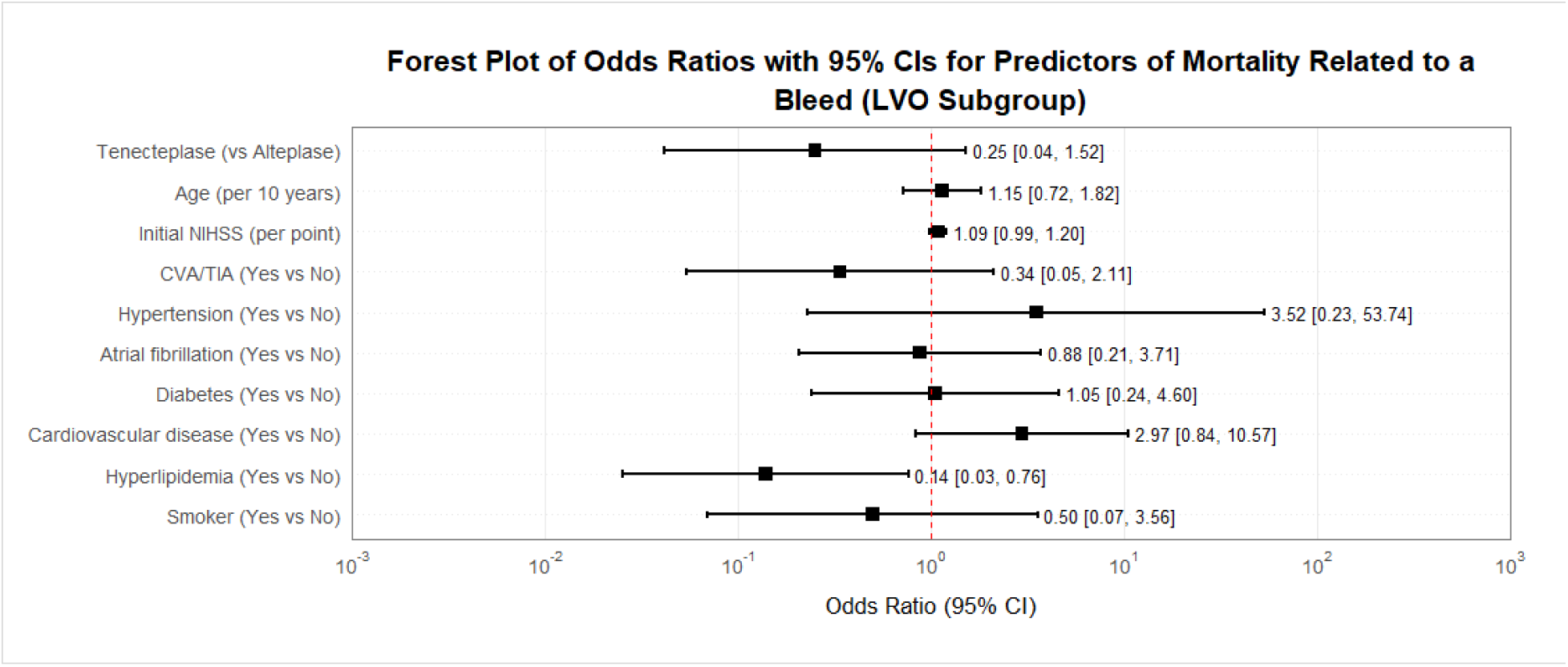
Adjusted odds ratios for predictors of mortality related to a bleed (LVO subgroup) based on binary logistic regression.

**Figure 8.**
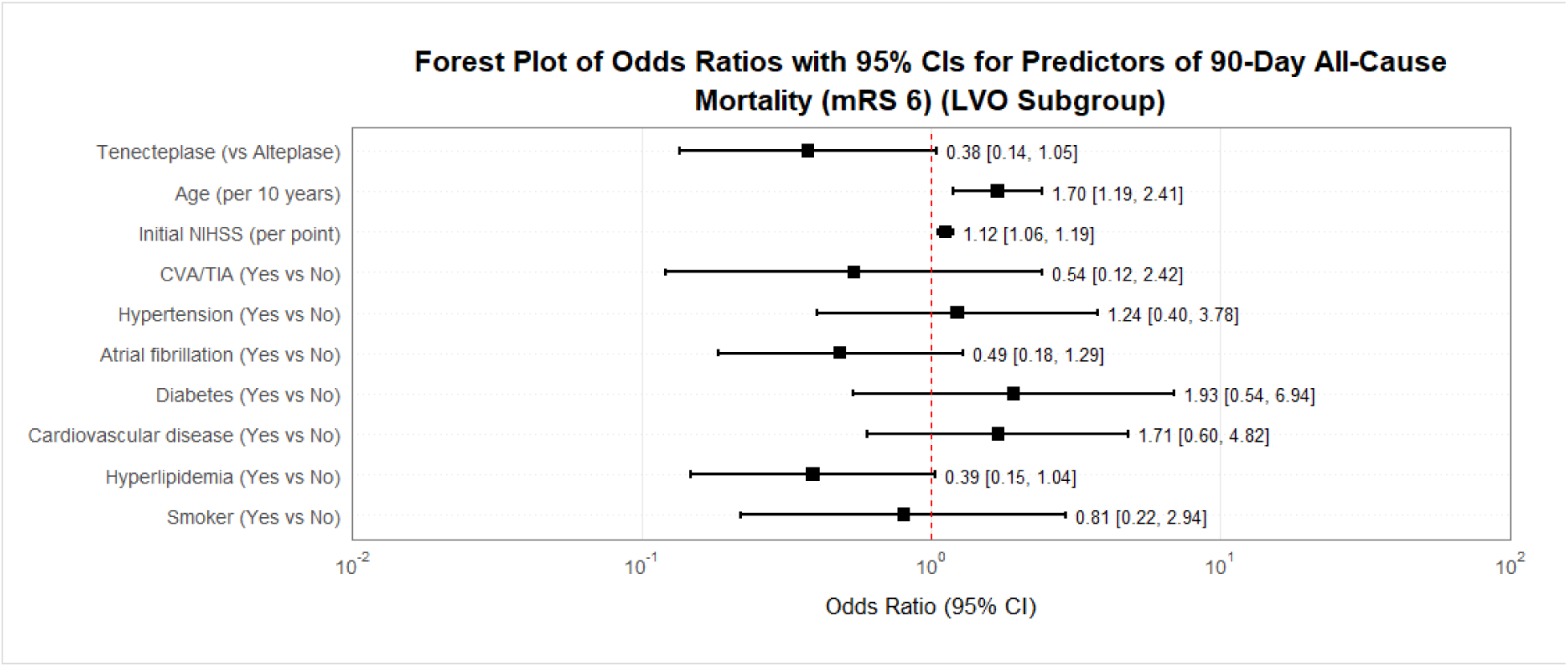
Adjusted odds ratios for predictors of 90-day all-cause mortality (mRS=6) (LVO subgroup) based on binary logistic regression.

**Figure 9.**
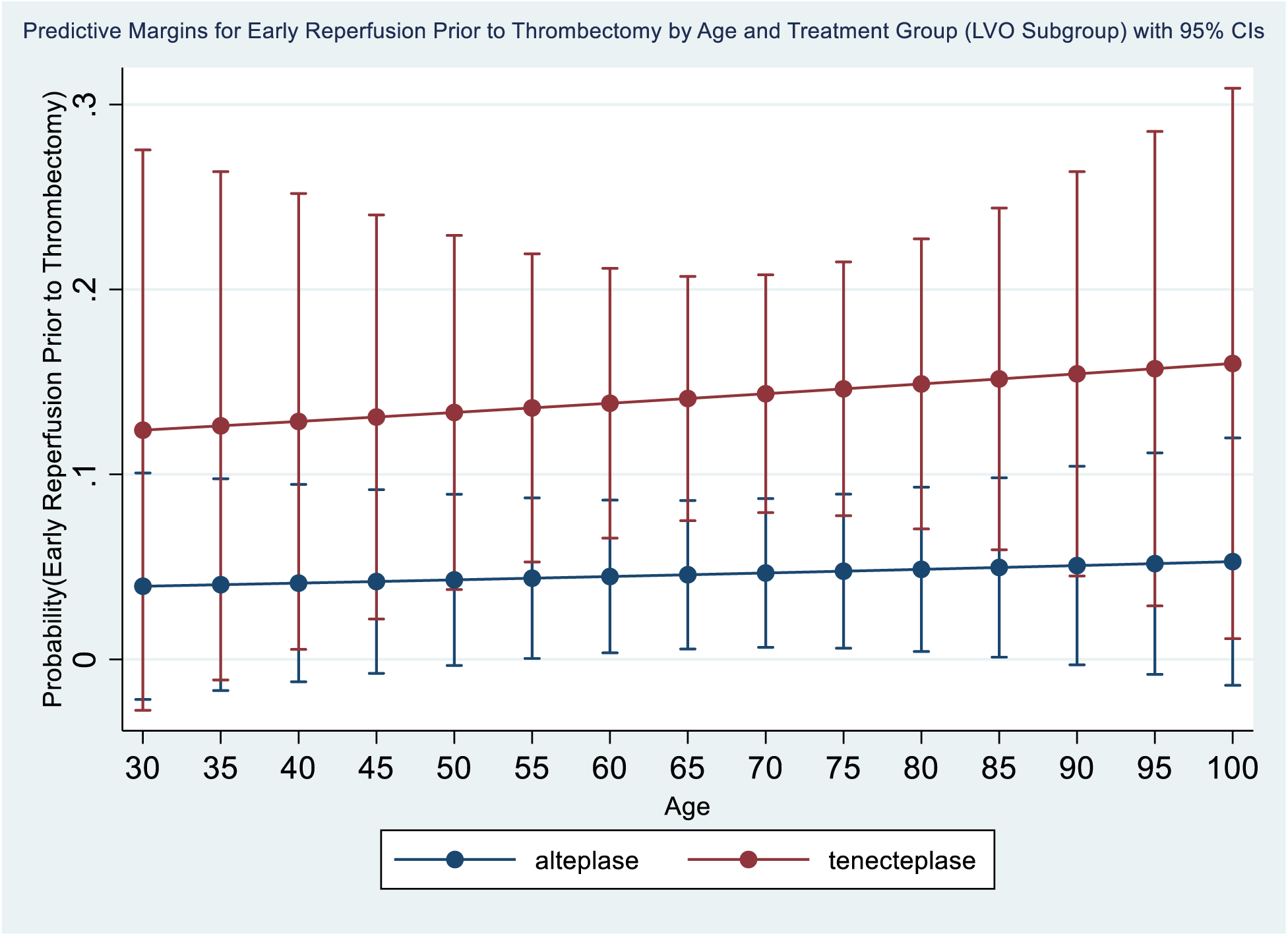
Predictive margins for early reperfusion prior to thrombectomy by age and treatment group (LVO subgroup) based on binary logistic regression.

**Figure 10.**
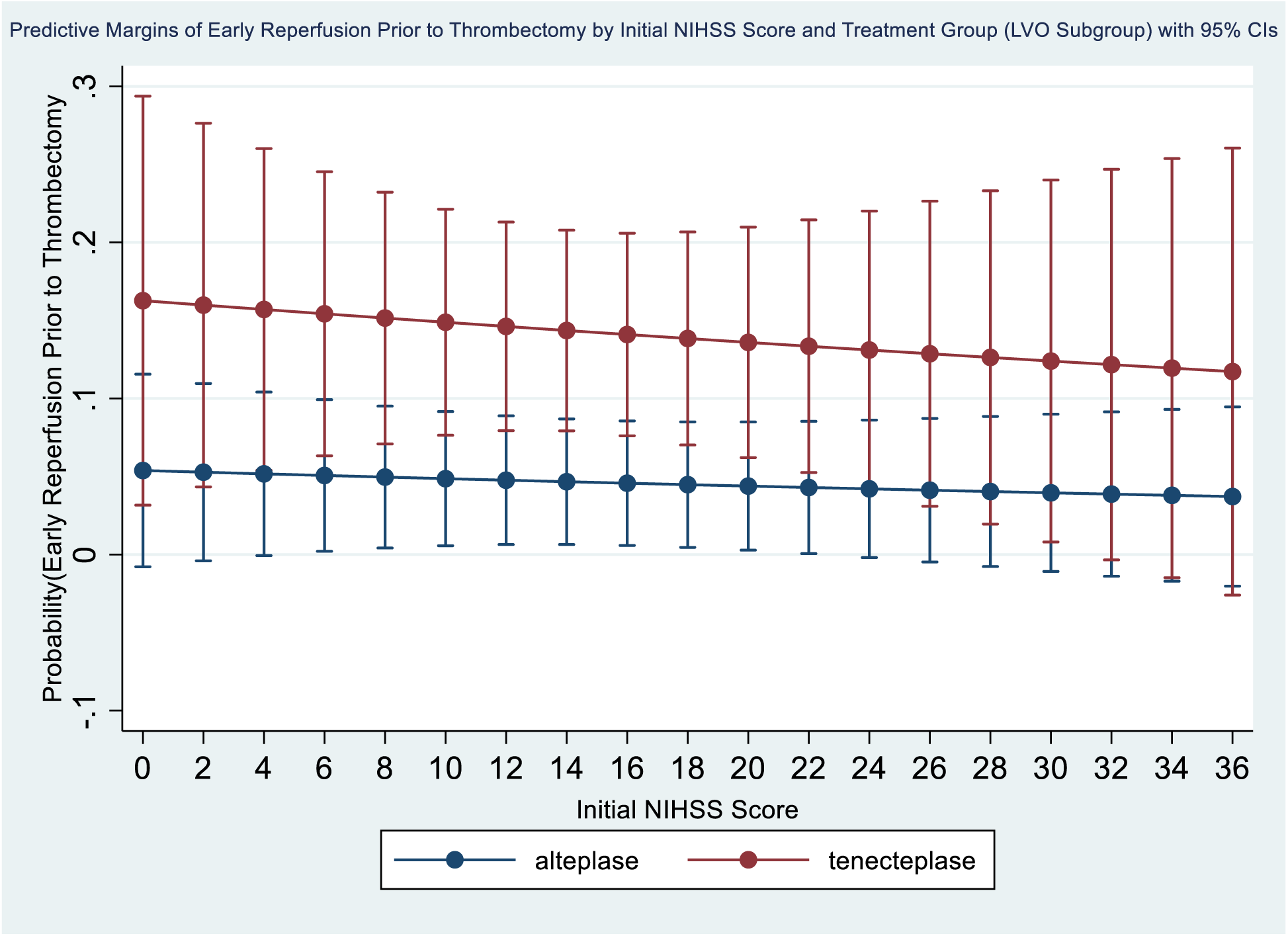
Predictive margins for early reperfusion without mechanical thrombectomy by initial NIHSS score and treatment group (LVO subgroup) based on binary logistic regression.

**Figure 11.**
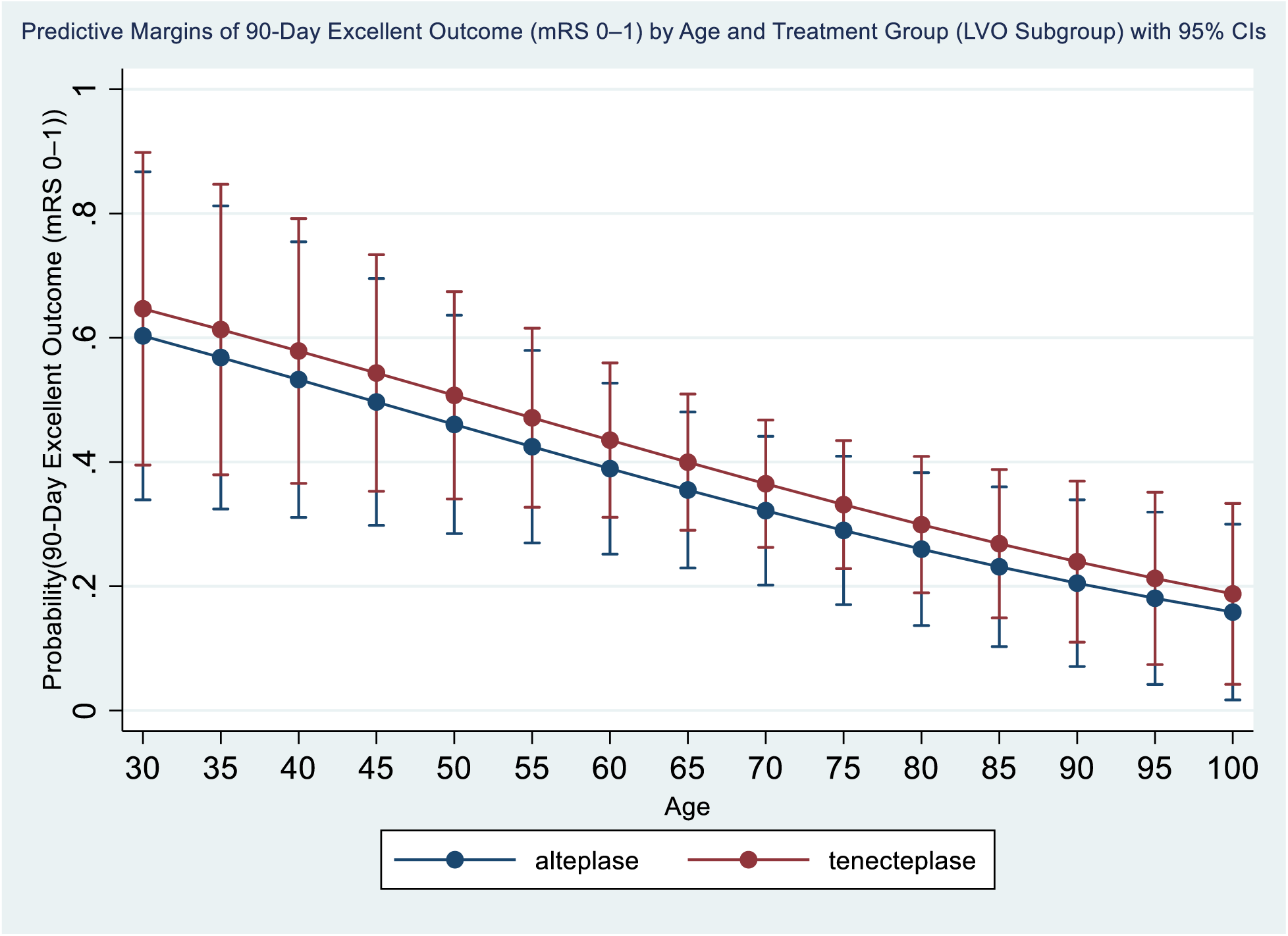
Predictive margins for 90-day excellent outcome (mRS 0–1) by age and treatment group (LVO subgroup) based on binary logistic regression.

**Figure 12.**
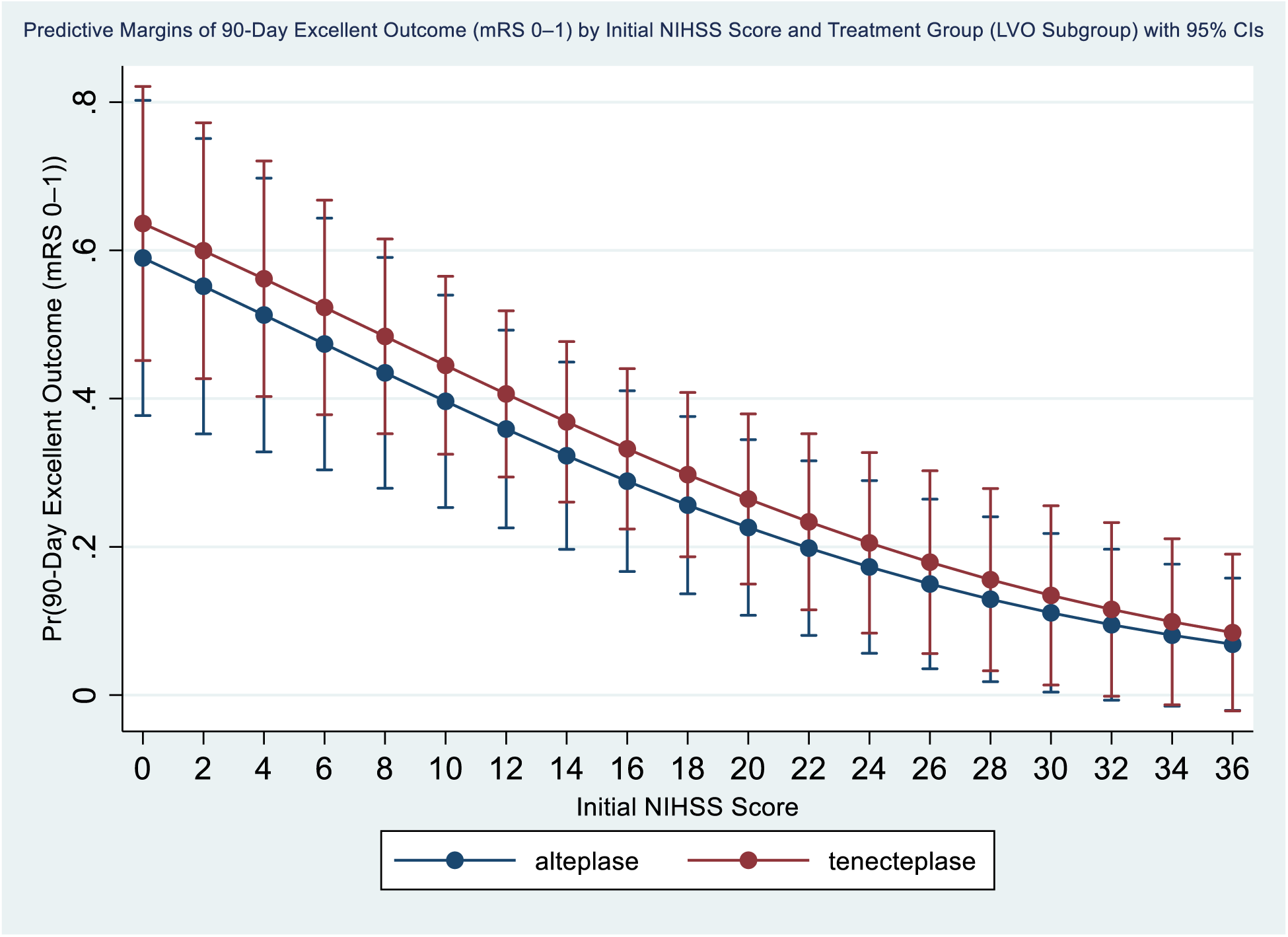
Predictive margins for 90-day excellent outcome (mRS 0–1) by initial NIHSS score and treatment group (LVO subgroup) based on binary logistic regression.

**Figure 13.**
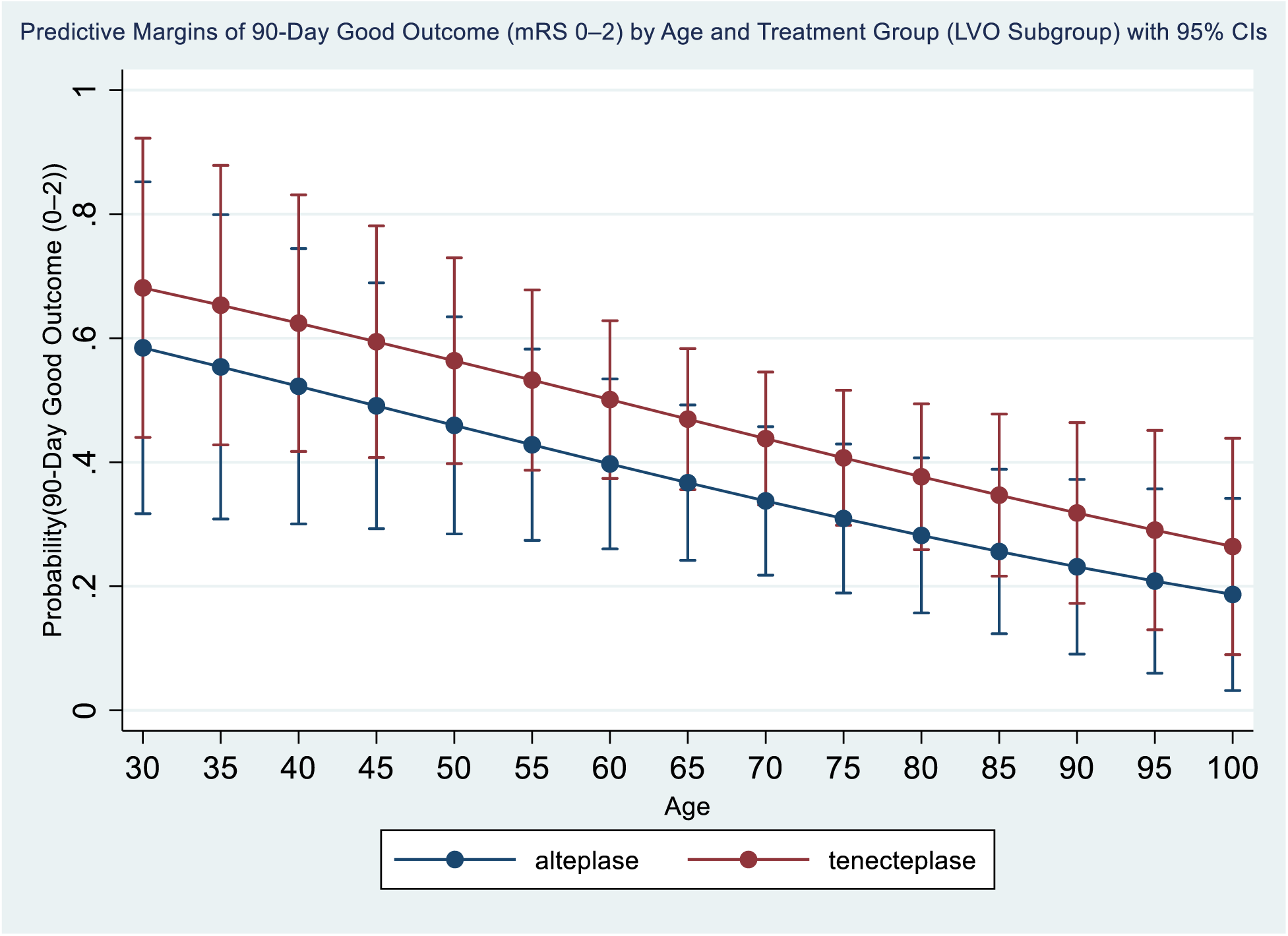
Predictive margins for 90-day good outcome (mRS 0–2) by age and treatment group (LVO subgroup) based on binary logistic regression.

**Figure 14.**
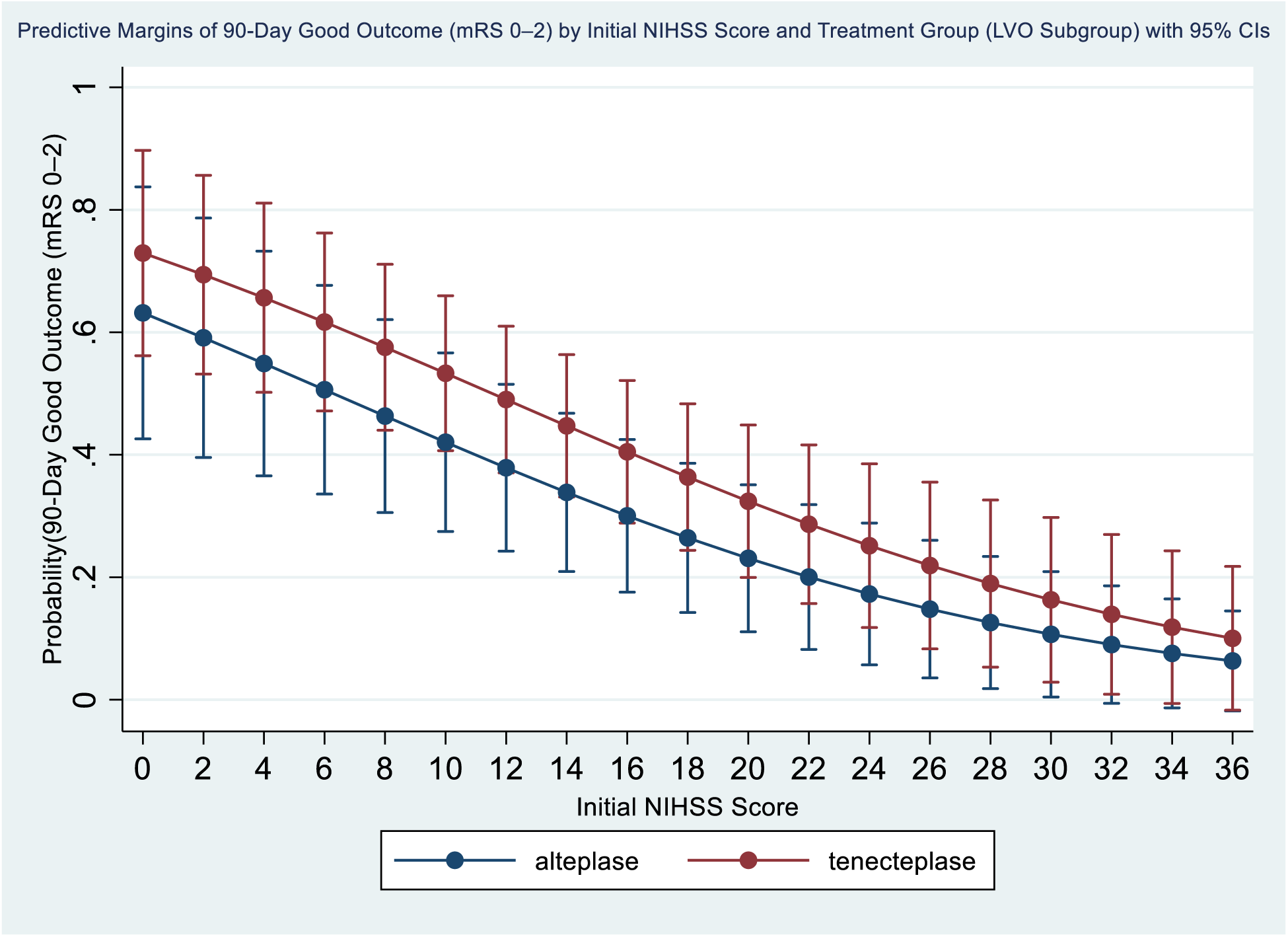
Predictive margins for 90-day good outcome (mRS 0–2) by initial NIHSS score and treatment group (LVO subgroup) based on binary logistic regression.

**Figure 15.**
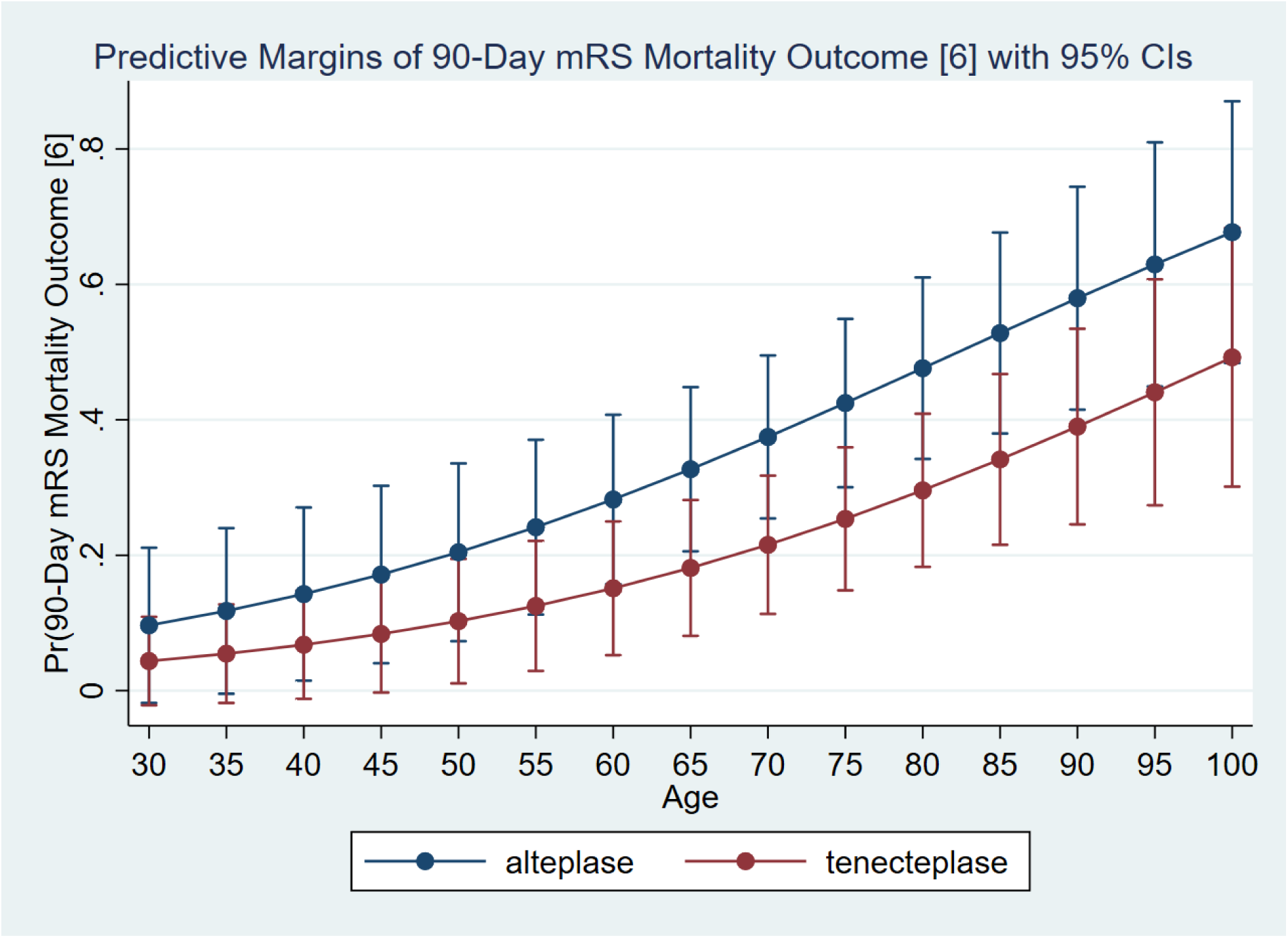
Predictive margins for 90-day mortality (mRS=6) by age and treatment group (LVO subgroup) based on binary logistic regression.

**Figure 16.**
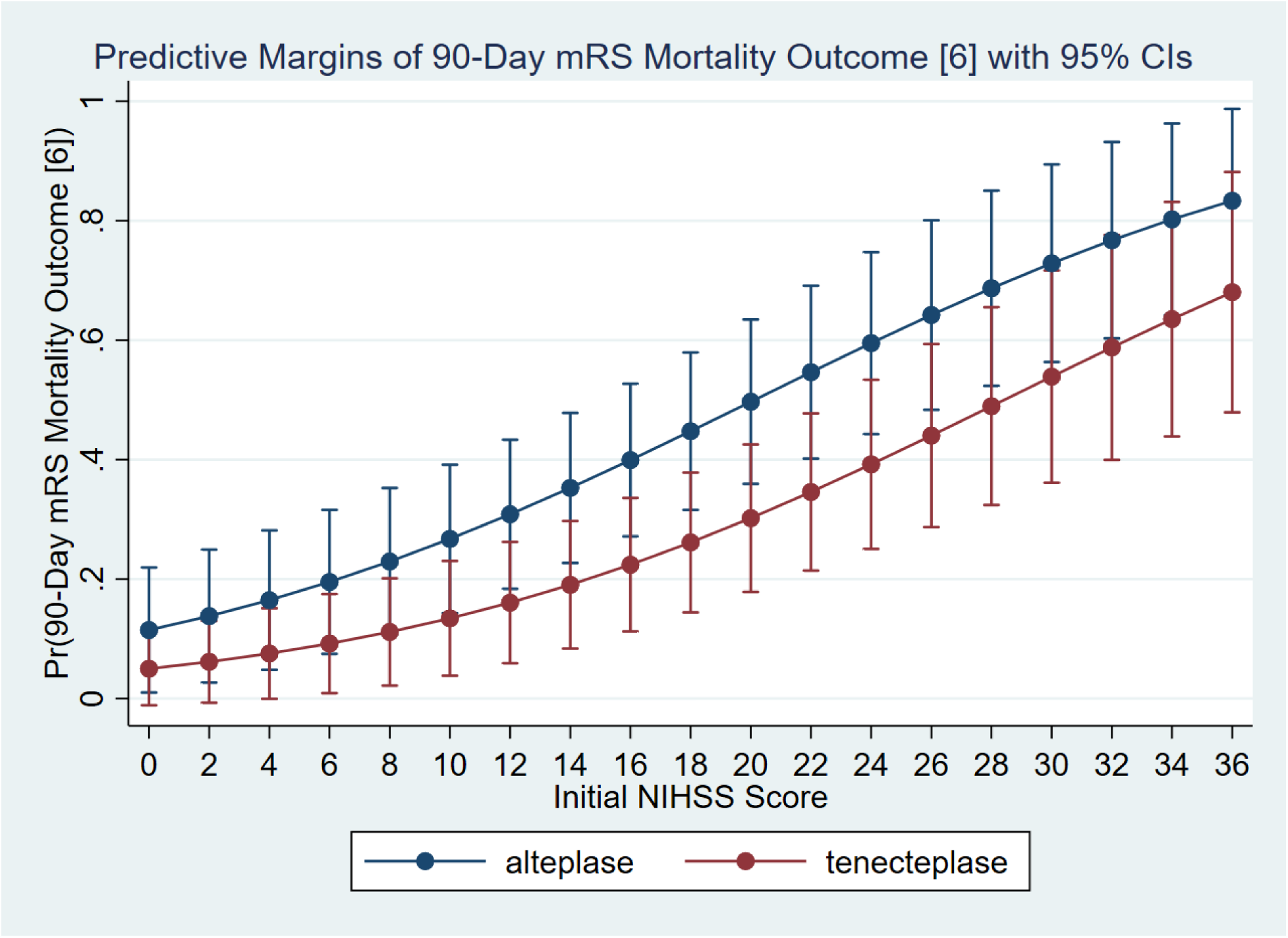
Predictive margins for 90-day mortality (mRS=6) by initial NIHSS score and treatment group (LVO subgroup) based on binary logistic regression.

## Appendix 1 Tenecteplase – (TNKase) Reconstitution and Dosing for Adult Stroke

Tenecteplase (TNKase) Dose:0.25 mg/kg (**not to exceed 25mg)** Give IV push over 5 seconds (Flush Line before and after with NS flush

- Open the Tenecteplase kit containing a 10 mL syringe, Red hub cannula filling device, TwinPak shield, Blunt plastic cannula, USP Sterile Water for Injection 10 mL, and 50 mg Tenecteplase vial.
- Use aseptic technique by washing hands and using gloves (Swab all vials with alcohol prep prior to reconstitution)
- Remove the flip-caps from 1 vial of Tenecteplase 50mg and 1 vial of 10ml Sterile Water for Injection.
- Flip the cap off both Sterile water for injection 10 ml vial and Tenecteplase vial. Use alcohol pads to wipe the surface of both vials.
- Reconstitution: Remove the syringe assembly from the syringe and set aside. Insert the syringe with the red cap cannula into the sterile water for injection vial. Aseptically withdraw all 10 mL from the SWFI vial into the syringe. Then inject the entire syringe into the Tenecteplase vial using the same red hub cannula directing the stream of water into the powder. Foaming may occur, which is normal. If there are any bubbles, let the vial stand for a minute until bubbles dissipate. **Swirl gently and do not shake**.
- Withdraw the appropriate volume based on chart below **dosed at 0.25 mg/kg with max dose of 25 mg**

### Administration of bolus (See Dosing Charts)

- **Step 1: Inspect Solution** - After reconstitution to concentration of mg/ml, inspect solution for particulate matter and discoloration.
- **Step 2: Administer Tenecteplase Dose -** Using a 10 ml Syringe give IV Push over 5 seconds (Follow the dosing chart for appropriate dosing)

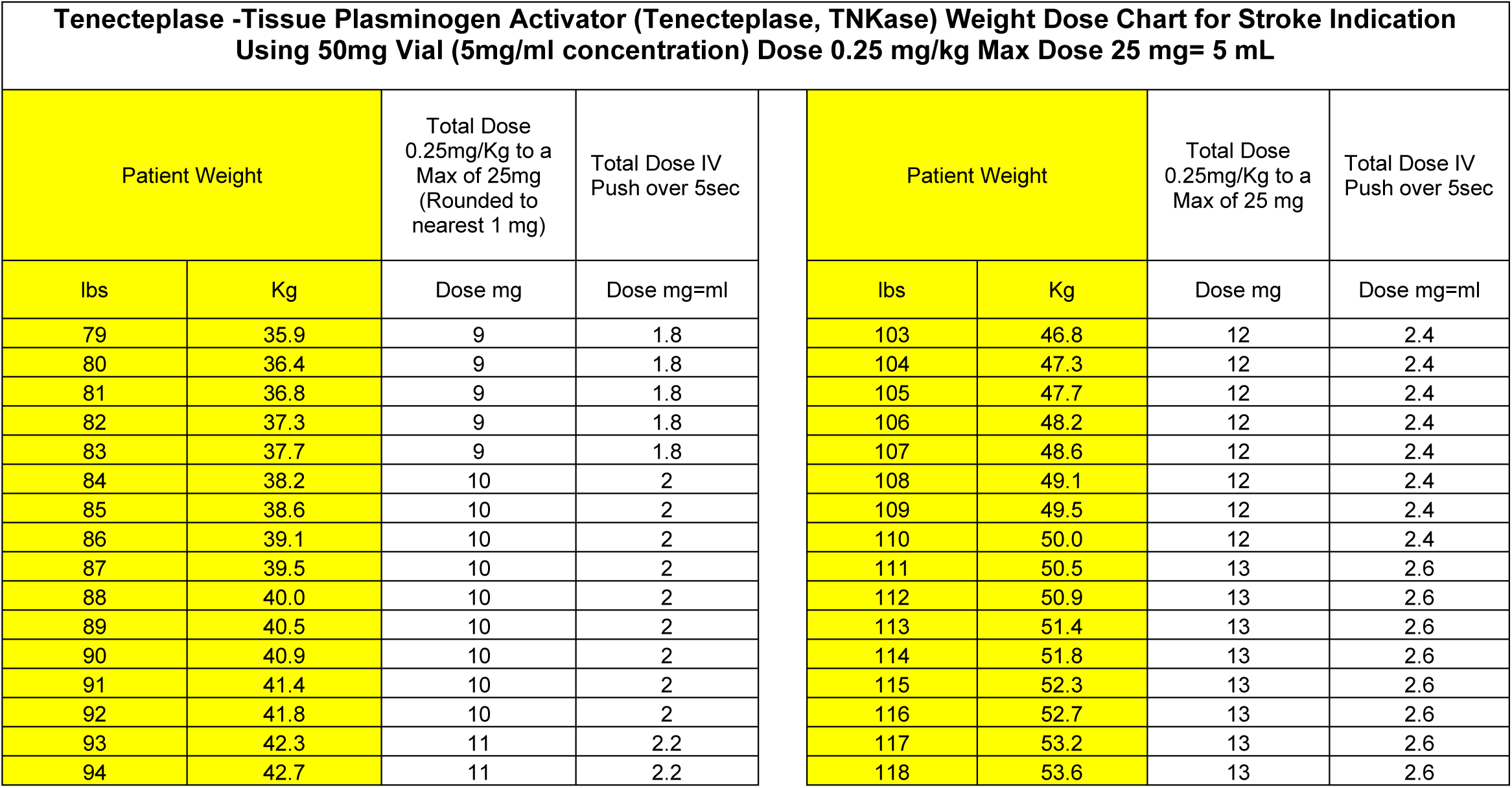
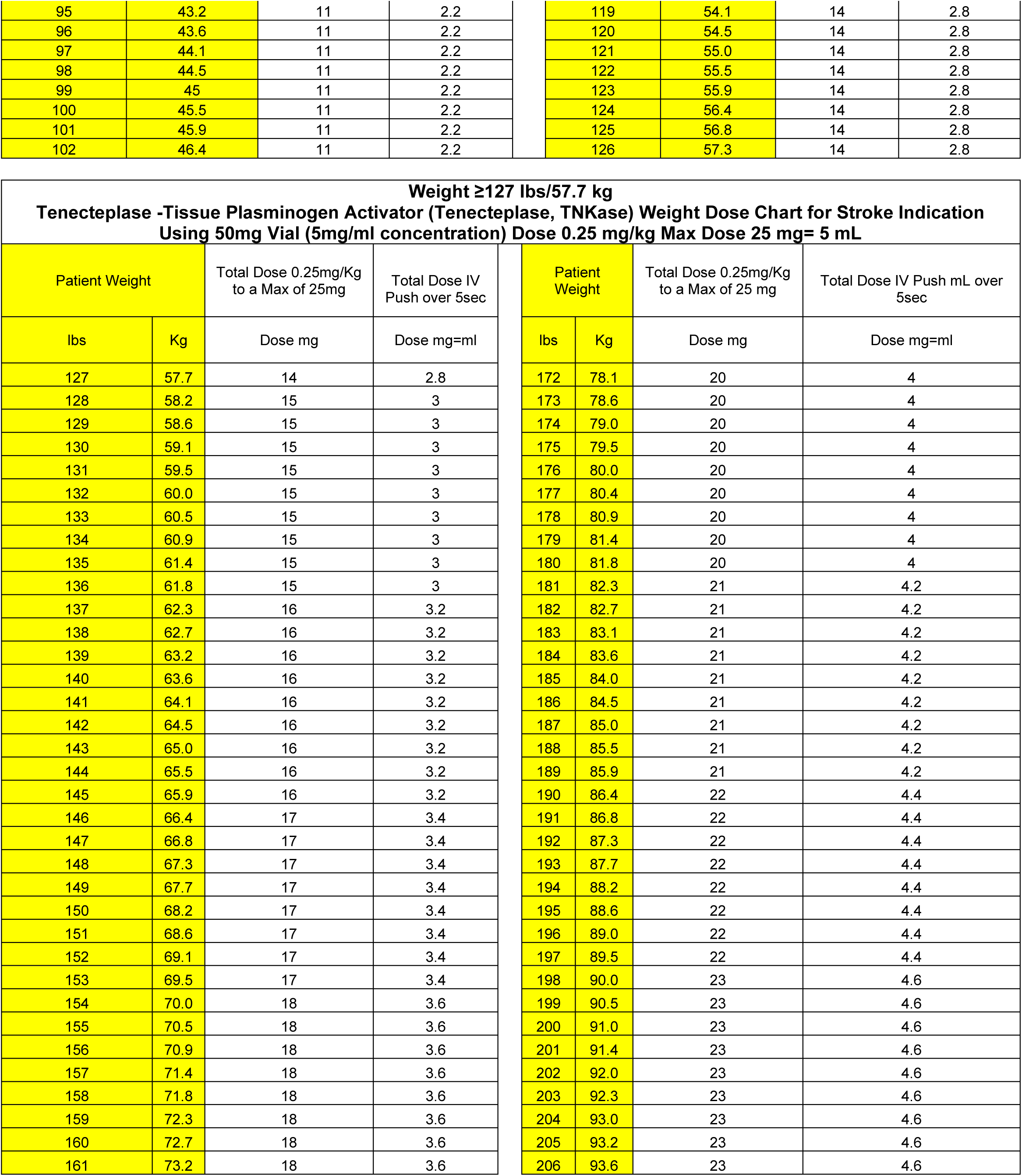
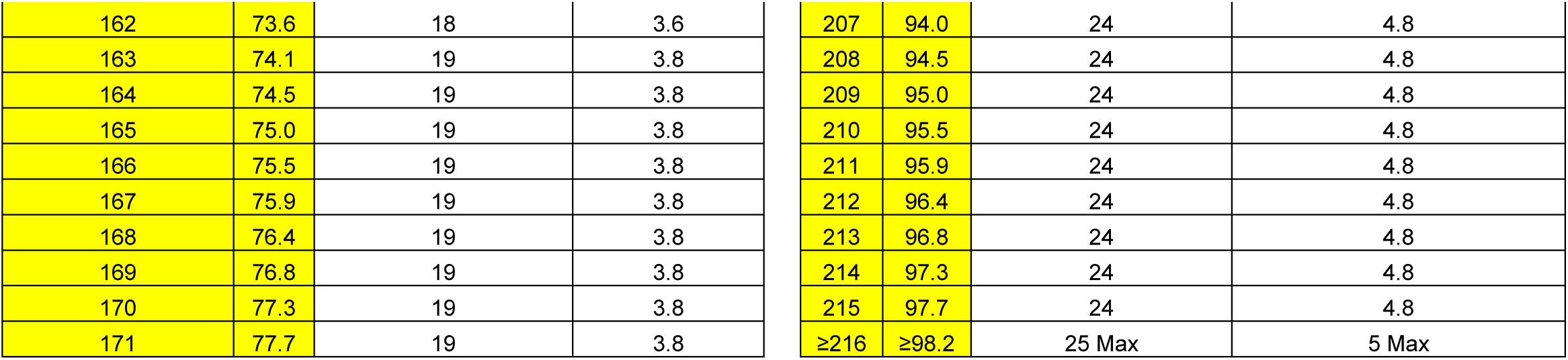

## Nonstandard Abbreviations and Acronyms

AIS: Acute Ischemic Stroke
TNK: Tenecteplase
LVO: Large Vessel Occlusion
DTN: Door-to-needle time
mRS: Modified Rankin Score
NIHSS: National Institute of Health Stroke
ICH: Intracerebral hemorrhage
NNT: Number Needed to Treat

